# Non-invasive brain stimulation effectiveness in anxiety disorder treatment: a meta-analysis on sham/behavior-controlled studies

**DOI:** 10.1101/2021.01.15.21249892

**Authors:** Alessandra Vergallito, Alessia Gallucci, Alberto Pisoni, Gabriele Caselli, Giovanni M. Ruggiero, Sandra Sassaroli, Leonor J. Romero Lauro

**Affiliations:** Department of Psychology, University of Milano Bicocca, Milano, Italy; Neuromi, School of Medicine and Surgery, University of Milano-Bicocca, Monza, Italy; Ph.D. Program in Neuroscience, School of Medicine and Surgery, University of Milano-Bicocca, Monza, Italy; Studi Cognitivi, Milan, Italy; Faculty of Psychology, Sigmund Freud University, Milan, Italy

**Author notes:** Corresponding author: Alberto Pisoni, PhD, Department of Psychology, University of Milano-Bicocca, Piazza dell’Ateneo Nuovo 1, 20126 Milano, Italy. AV was supported in part by a 2019 NARSAD Young Investigator Grant from the Brain & Behavior Research Foundation.

**Keywords:** Non-invasive brain stimulation, rTMS, tDCS, Anxiety disorders, Specific phobia, Social anxiety disorder, Panic disorder, Agoraphobia, Generalized anxiety disorder

## Abstract

**Background:** Recently, the possibility of using non-invasive brain stimulation (NIBS) to treat mental disorders received considerable attention. Repetitive transcranial magnetic stimulation (rTMS) and transcranial direct current stimulation (tDCS) are considered effective treatments for depressive symptoms. However, no recommendation is available for anxiety disorders, suggesting that evidence is still limited.

**Objective:** We systematically revised the existing literature, and quantitatively analyzed the effectiveness of rTMS and tDCS in anxiety disorders treatment.

**Method:** Following PRISMA guidelines, 3 electronic databases were screened to the end of February 2020 to select English-written peer-reviewed articles including (i) a clinical sample of patients with anxiety disorders, (ii) the use of a NIBS technique, (iii) the inclusion of a control condition, and (iv) pre-post scores at a validated questionnaire measuring anxious symptoms.

**Results:** Eleven papers met the inclusion criteria, comprising 154 participants assigned to the real stimulation condition and 164 to the sham or control group. The *Hedge g* for scores at disorder specific and general anxiety questionnaires before and after the treatment was computed as effect size and analyzed in two independent random-effects meta-analyses. Considering the well-known comorbidity between anxiety and depression, a third meta-analysis was run, analyzing depression scores outcomes. Results showed a significant effect of NIBS in reducing questionnaires scores in the real vs. control condition at specific and general anxiety measures, and depressive symptoms.

**Conclusion:** Albeit preliminary, our findings highlighted that real stimulation reduced anxiety and depression scores compared to the control condition, suggesting that NIBS can alleviate clinical symptoms in patients with anxiety diseases.

## Introduction

Anxiety disorders are the most prevalent class of mental disorders in most western societies ^1, 2^ and are one of the foremost causes of disability ^3^ (for epidemiologic details see ^4^). The onset of anxiety disorders typically occurs within young adulthood ^5^. It seems to take a chronic course, with differences in stability rates varying across studies and specific diagnosis ^6^.

According to the Diagnostic and Statistical Manual of Mental Disorders (DSM), Fifth Edition ^7^, the anxiety disorders category includes specific phobia, social anxiety disorder, panic disorder, agoraphobia, and generalized anxiety disorder^1^. DSM-5 anxiety diagnostic criteria are similar to those included by the other standard classification system, namely the International Classification of Diseases, Tenth Edition, or ICD-10 ^8^. Across the two systems, anxiety disorders are a spectrum of multidimensional phenotypes ^9^, that share clinical features, such as excessive and stable anxiety, physiological symptoms, such tachycardia, and chest tightness, typical behavioral responses, such as avoiding perceived threats, places or situations, thus impairing individuals’ psychological well-being and quality of life.

The neurobiology of anxiety disorders is far from being clarified. Indeed, studies have been conducted over small participant samples, with heterogeneous imaging methods, paradigms, and patients’ comorbidities ^10^.

Despite disease-specific differences, converging evidence suggests that anxiety disorders are characterized by structural and functional alterations, primarily involving a meso cortico limbic pathway (see for a review ^11^). According to this neurobiological account, the amygdala, the prefrontal cortex (PFC), the anterior cingulate cortex (ACC), the hippocampus, and their functional connections, might play a key role in generating and regulating fear, anxiety, and threat detection ^4, 9^. Amygdala hyperactivity is one of the most consistent findings ^12, 13^. This abnormal activity has been reported across several specific diseases and tasks, such as anxiety-provoking public speaking ^14–16^, fear-conditioning ^17^, or emotional images/threatening faces presentation tasks in social phobia or generalized social anxiety disorder ^18, 19^. Moreover, amygdala activation positively correlated with symptoms’ severity ^20, 21^ and decreased after medication and psychotherapy interventions ^21–24^. Amygdala response to threat is regulated through bidirectional connections to the ACC and vmPFC in animals and humans ^25, 26^. In line with this finding, human neuroimaging studies highlighted PFC hypoactivity in anxious patients, suggesting that amygdala hyperactivity might be due to a decrease in the top-down inhibitory control exerted by the prefrontal cortex ^27–30^ (but see ^31^ for different results). Considering functional abnormalities in anxiety disorders, it has been suggested that an inter-hemispheric imbalance might be at the basis of the disease, involving a hypoactivation of the left dorsolateral prefrontal cortex (DLPFC) and a hyperactivation of the right DLPFC ^32–34^.

Anxiety first-line treatments comprise pharmacological or psychotherapeutic interventions, with Cognitive-Behavioral therapy considered as the most effective treatment according to several international guidelines ^35, 36^. However, a consistent number of patients do not respond to traditional treatment or go through relapse and recurrence of their symptoms ^37, 38^.

In the search for alternative treatments, in the last thirty years, the use of non-invasive brain stimulation (NIBS) has rapidly grown both as a stand-alone therapy or combined with cognitive or behavioral interventions ^39–41^. NIBS’ rationale in psychiatric treatment is the possibility of re-balancing maladaptive activity and functional connectivity between brain structures, which can be altered in these disorders. Indeed, there is a consensus that aside genetic, hormonal, social, and cognitive factors, psychiatric disorders involve also pathologically altered neural plasticity, which can be modulated through NIBS with biochemical effects that outlast the time of stimulation ^42^ (see for recent reviews ^43, 44^). Although their precise action mechanisms are still under investigation, NIBS effects on synaptic plasticity involve several phenomena ultimately leading to long-term potentiation (LTP) – or synaptic strengthening – and long-term depression (LTD) – or synaptic weakening – processes ^45^ (for a review and discussion see ^46^).

Among NIBS techniques, the two most used methods are transcranial magnetic stimulation (TMS) and transcranial direct current stimulation (tDCS). TMS is a technique based on delivering a strong, short magnetic pulse to the participants’ head, inducing neuronal firing by suprathreshold neuronal membrane depolarization ^47^. When used to generate long-term effects, TMS is typically applied using repetitive (rTMS) protocols, with inhibitory (≤1 Hz and continuous theta burst stimulation, cTBS), or excitatory (higher than 5 Hz and intermittent theta burst stimulation, iTBS) protocols ^48^. In turn, tDCS is a neuromodulatory technique in which weak constant direct current (typically 1-2 mA) is delivered through the scalp using two electrodes, one with positive (anode) and the other with negative (cathode) polarity ^49^. tDCS does not generate action potentials per se but induces small changes at the membrane potential level, thus influencing spike frequency and, in turn, cortical excitability ^50, 51^. TDCS effects are polarity-dependent, with anodal stimulation depolarizing neuronal membrane and cathodal hyperpolarizing it, increasing, and decreasing cortical excitability, respectively ^52^. Usually, among the NIBS parameters, the stimulation frequency for TMS (either high or low) and the polarity for tDCS (either anodal or cathodal) are considered the determinants for an expected effect in cortical excitability and behavior: excitatory-enhancing effect or inhibitory-disrupting effect. Although a detailed discussion of the two techniques goes beyond this meta-analysis scope, it seems crucial to highlight that such expectation can be misleading. Indeed, the NIBS outcome -in terms of both cortical excitability and behavioral modulation - can’t be clearly determined in advance but is the result of more complex interactions involving stimulation parameters (intensity, orientation), cerebral regions and their connections, individual anatomical features, and state dependency ^53–56^.

Among psychiatric disorders, the main field of application of NIBS as an alternative treatment is the major depression disorder (MDD). rTMS clinical use to treat MDD has been approved by the Food and Drug Administration in 2008 using high-frequency (10 Hz) left-side dorsolateral prefrontal cortex stimulation, and in 2018 applying iTBS protocols over the same region ^57^. The effectiveness of rTMS and tDCS in other psychiatric disorders has been explored by several reviews and meta-analyses, targeting schizophrenia ^58, 59^, substance abuse ^60^, and obsessive-compulsive disorders ^61, 62^, highlighting promising yet preliminary results.

Aiming at providing shared recommendations for good practice, periodically revised guidelines of independent expert panels review and analyze studies investigating rTMS ^63, 64^ and tDCS ^65, 66^ protocols applied to a broad spectrum of neurological and psychiatric disorders. According to the guidelines’ classification, Level A (“definitely effective or ineffective”) indicates that evidence is sufficient (in terms of number and studies quality) to establish whether a specific protocol applied over a certain region can be useful or not in a particular disorder. Only a few protocols reached level A of efficacy. Specifically, for TMS Level A effectiveness is attributed to i) high frequency rTMS applied to the left DLPFC to treat depression; ii) high frequency TMS to the primary motor cortex (M1) contralateral to the painful side for neuropathic pain; iii) low frequency rTMS applied over the contralesional M1 for hand motor recovery in the post-acute stage of stroke ^64^. Concerning tDCS, level A effectiveness is attributed only to anodal stimulation over the left DLPFC in depression ^65^, while anodal tDCS over the ipsilesional primary motor cortex is considered definitely not effective for enhancing robotic therapy in the motor rehabilitation in subacute stroke. Critically, to date, no recommendation has been made for the use of rTMS or tDCS in anxiety disorders treatment. Indeed, the available data are not sufficient to make recommendations to its use or a claim for an absence of an effect ^64, 65^.

Trying to fill the gap in the literature, in the last few years, several reviews examined the available literature concerning the therapeutic effects of rTMS and tDCS in anxiety disorders treatment ^34, 67^, anxiety symptoms arising from other pathologies ^68^, and specific anxiety disorders (e.g., generalized anxiety disorder ^69^). These works testified the general interest around this topic and showed promising yet preliminary results. However, so far, previous reviews also included single-case studies and protocols without a control condition, thus providing an overview of the literature state-of-the-art, but without cumulatively quantifying the results.

To our knowledge, three previous meta-analyses ^70–72^ investigated rTMS efficacy from a quantitative perspective. Specifically, Cui and colleagues investigated rTMS efficacy in treating GAD. The meta-analysis included 21 studies (2 English-written and 19 Chinese), with the inclusion criteria of a control group receiving sham rTMS or no intervention, suggesting rTMS as a useful option in decreasing GAD anxiety symptoms. The review and meta-analysis by Trevizol and colleagues ^72^ investigated rTMS efficacy in randomized clinical trials addressing anxiety disorders. This work included 14 papers; however, five works addressed post-traumatic stress disorder and eight obsessive-compulsive disorders, which are now considered as independent diagnostic categories ^7^. The authors concluded that real TMS was not superior to the sham condition in reducing anxiety symptoms. In line with the previous study, Cirillo and colleagues ^70^ systematically reviewed and analyzed anxiety and posttraumatic stress disorders (PTSD), including 17 papers: nine concerning PTSD, two specific phobias, two addressing panic disorders and four GAD. Authors then run two independent meta-analyses, one concerning PTSD and the second including GAD. Authors considered the mean difference of sham vs. real TMS in pre- and post-treatment scores when the two conditions were available, only pre-post scores mean difference when sham stimulation was not tested. Results showed a large treatment efficacy for both disorders.

To our knowledge, no previous meta-analyses combined TMS and tDCS in investigating NIBS effectiveness in treating anxiety disorders. Moreover, some of the previous works included research not involving a control group or considering disorders that are now considered as separated nosological entities. Therefore, the present work aims to qualitatively visualize and quantitatively evaluate the effect induced by ether rTMS and tDCS protocols in anxiety disorders, trying to overcome the limitation of individual studies which have been typically conducted on small sample sizes and applying heterogeneous stimulation parameters and sessions number ^34^.

Since a similar neurobiological pattern of imbalance of cortical excitability between right and left DLPFC has been reported in major depression disorder ^73–75^, which is in line with the frequent comorbidity of the two disorders ^76–78^, studies in which anxious patients had a comorbid depression diagnosis were included in our meta-analysis. Indeed, despite anxiety and depression have been considered two nosologically independent categories according to the traditional classifications, their comorbidity is well-known in clinical practice and research (for a recent revision on comorbid anxiety and depression syndrome, see ^79^).

## Methods

The Preferred Reporting Items for Systematic Reviews and Meta-Analyses (PRISMA) guidelines ^80, 81^ were used to conduct this systematic review and meta-analysis.

### Literature search

PubMed, Web of Science, and Scopus were used to select peer-reviewed original papers published in English and before the end of February 2020, exploring the application of rTMS or tDCS in samples of patients suffering from anxiety-related disorders. We combined keywords related to the brain stimulation techniques “rTMS”, “tDCS” with the relevant anxiety disorders labels “generalized anxiety disorder”, “agoraphobia”, “panic disorder”, “specific phobia”, “social anxiety”. We excluded non-English written papers, case reports, systematic and narrative reviews, meta-analyses, conference proceedings, and abstracts. Papers measuring anxiety within non–clinical populations, without a sham or behavioral controlled condition and at least one clinical validated questionnaire were also excluded. Moreover, when multiple papers were based on the same dataset, we included only the first paper reporting the relevant results on the clinical sample.

### Records screening and data extraction

To blind the screening process, we used Rayyan (https://rayyan.qcri.org/), a web and mobile systematic reviews manager ^82^. After removing duplicates, three researchers (A.V., A.G., A.P.) independently filtered the records as “include”, “exclude”, or “maybe” considering publications’ titles and abstracts. Reasons for exclusion were specified through defined labels based on the inclusion and exclusion criteria. Then, the full text of the remaining records was analyzed by three researchers (A.V., A.G., A.P.), who independently selected the eligible studies. When the articles’ full versions were not available, the corresponding authors of the papers were contacted. In both the title/abstract and full-text screening phase, conflicting decisions were solved by consensus. One researcher (A.P.) extracted data by using a previously prepared structured form (Table 1), checked for consistency and accuracy by other two authors (A.V., A.G.). Discrepancies were resolved by agreement of three authors (A.V., A.G., A.P.).

**Table 1.**
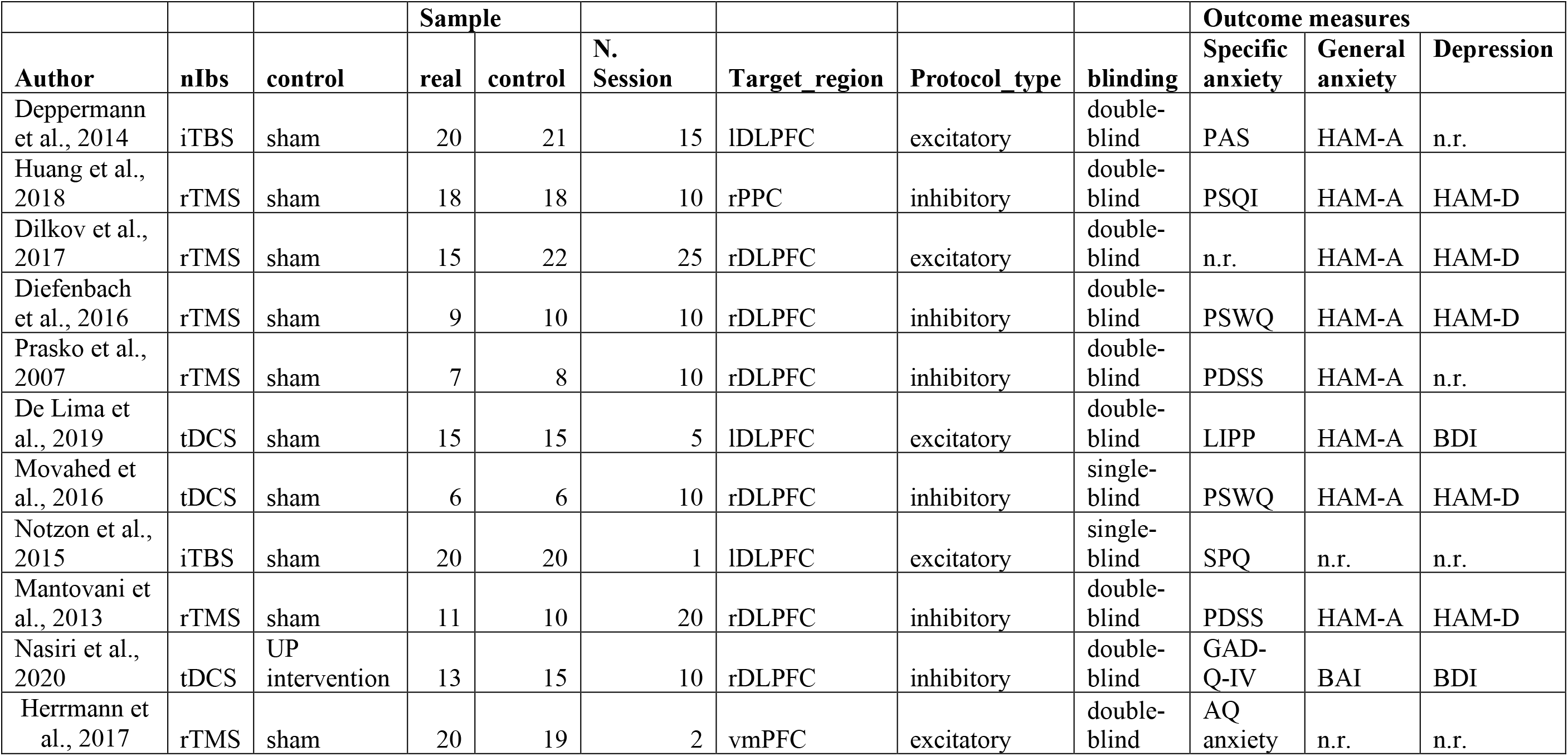
summarizes studies’ features used for quantitative analysis. iTBS = intermittent theta burst stimulation; rTMS = repetitive transcranial magnetic stimulation; tDCS = transcranial direct current stimulation; lDLPFC=left dorsolateral prefrontal cortex; rDLPFC = right dorsolateral prefrontal cortex; vmPFC = ventromedial prefrontal cortex; PAS = Panic and Agoraphobia Scale; PSQI = Pittsburgh Sleep Quality Index; PSWQ = Penn State Worry Questionnaire; PDSS = Panic Disorder Severity Scale; LIPP = Lipp Inventory of Stress Symptoms for Adults; SPQ = Spider Phobia Questionnaire; GAD-Q-IV = Generalized Anxiety Disorder Questionnaire-IV; AQ anxiety= Acrophobia Questionnaire anxiety subscale; HAM-A = Hamilton Anxiety Rating Scale; HAM-D = Hamilton Rating Scale for Depression; BDI = Beck Depression Inventory; BAI = Beck Anxiety Inventory; n.r. = not reported.

Measures of anxiety levels were extracted from the selected studies, namely a more “general” one, typically consisting of the Hamilton Anxiety Rating Scale ^83^ (HAM-A or HAR S) or the Beck Anxiety Interview ^84^ (BAI) and, when present, a disorder-specific one, which changed depending on the specific disorders (e.g., the Panic Disorder Severity Scale, PDSS ^85^ when participants were recruited based on panic disorder, see the outcome measure paragraph for details). Moreover, given the high comorbidity of anxiety and depressive disorders, when available, depression measures, the Hamilton Depression Rating Scale ^86^ (HAM-D or HDRS) or the Beck Depression Inventory ^87^ (BDI) were also collected and analyzed.

### Qualitative analysis

Two researchers (A.V., A.G.) independently assessed the quality of the studies retrieved based on the following Cochrane Collaboration’s Risk-of-Bias Tool ^88^ criteria: random sequence generation, allocation concealment, blinding strategy, incomplete outcome data, selective outcome reporting. For the selection bias, the random sequence generation item was rated as “low risk” only when the randomization procedures (e.g., random number table, computer-generated randomization, randomization envelopes) were reported. The allocation concealment item was rated as “low risk” only for studies that recruited a group of patients who received sham stimulation. For the reporting bias, when available, we checked the registered protocol of the included records. Conflicts were solved by consensus of the two researchers and by consulting a third researcher when needed (A.P.).

### Quantitative analysis

For each included study, relevant information was extracted, including means, standard deviations of the scores at the clinical scales, NIBS protocol (technique, number of sessions, stimulation location), and patients characteristics. As the primary outcome measure, the pre-post treatment mean difference in specific scales for anxiety disorders (9 studies included, see the outcome measures paragraph for details) was extracted for the experimental and the control group to measure the impact of the NIBS protocol over the anxiety symptoms. When sufficient information was not reported in the articles’ text, tables, or supplementary material, the authors were contacted to obtain these missing data ^89–91^. The standard deviation of the change score (pre- to post-NIBS treatment) was calculated, as suggested by the Cochrane handbook for systematic reviews of interventions ^92^, as:

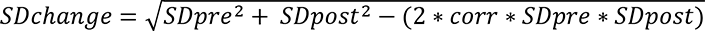

where *corr* is the correlation between pre- and post-measurements variances, set at .5 as suggested by Follman and colleagues ^93^.

Sampling variance, standardized mean difference (SMD), and summary analyses were computed for each included study, using the “escalc” function of “metafor” package for R, version 3.4.3 ^94^. SMD was corrected for the positive bias for small group within the function, computing Hedge’s g ^95^, which was used as an effect size measure. In this meta-analysis, we computed the global effect of NIBS protocol in reducing anxiety symptoms using a random-effects model with the “rma” function of the “metaphor” R package regardless of the heterogeneity tests values: studies, indeed, vary according to their characteristics (e.g., patients’ characteristics, stimulation interventions, associated therapies, etc.). Nevertheless, heterogeneity was assessed through both the variation due to the sampling error (Q statistic) and the percentage of variation between studies due to heterogeneity rather than chance (I^2^ statistics) ^96^. Potential outliers were identified with the analysis of influence ^97–99^, implemented in the “inf” function of the “metaphor” R package. Finally, publication bias was controlled for with the funnel plot, the Egger’s regression test ^100^ and the rank correlation test ^101^, and eventually corrected with the “trim and fill” method ^102^, which creates dummy potential missing studies to create a more symmetrical funnel plot. Finally, we run a moderation analysis to test whether the duration of the treatment (computed in the number of stimulation sessions), the stimulation technique applied (iTBS, rTMS, tDCS), the protocol type (excitatory vs. inhibitory), the target region^2^ (left vs. right DLPFC) and comorbid depression (presence vs. absence) influenced the obtained effects.

The same procedures were adopted for the secondary outcome measures, namely the general anxiety scale (HAM-A or BAI, nine studies included) and depressive interview/self-report questionnaire (HAM-D or BDI, seven studies included).

## Results

### Studies selection

Eight hundred seventy-six publications were identified. We removed duplicates and carefully reviewed the title and abstract of the remaining 637 records. Among these, 611 were excluded since the inclusion criteria were not met, while the full texts of the remaining 26 papers were examined. Eleven studies met the qualitative and quantitative analysis inclusion criteria. Six studies were excluded because they did not consider a control condition, namely sham stimulation, or a control group ^103–108^. Five papers were excluded because they involved samples already analyzed in previous articles ^109–113^. One paper was excluded because it did not include patients ^28^, one included patients but without a diagnosis of an anxiety-related disorder (Caulfield & Stern, 2019), and one did not test anxiety as an outcome measure ^115^. Finally, one paper was excluded because the full text was not available ^116^. Table 2 summarizes the selection procedure.

**Table 2.**
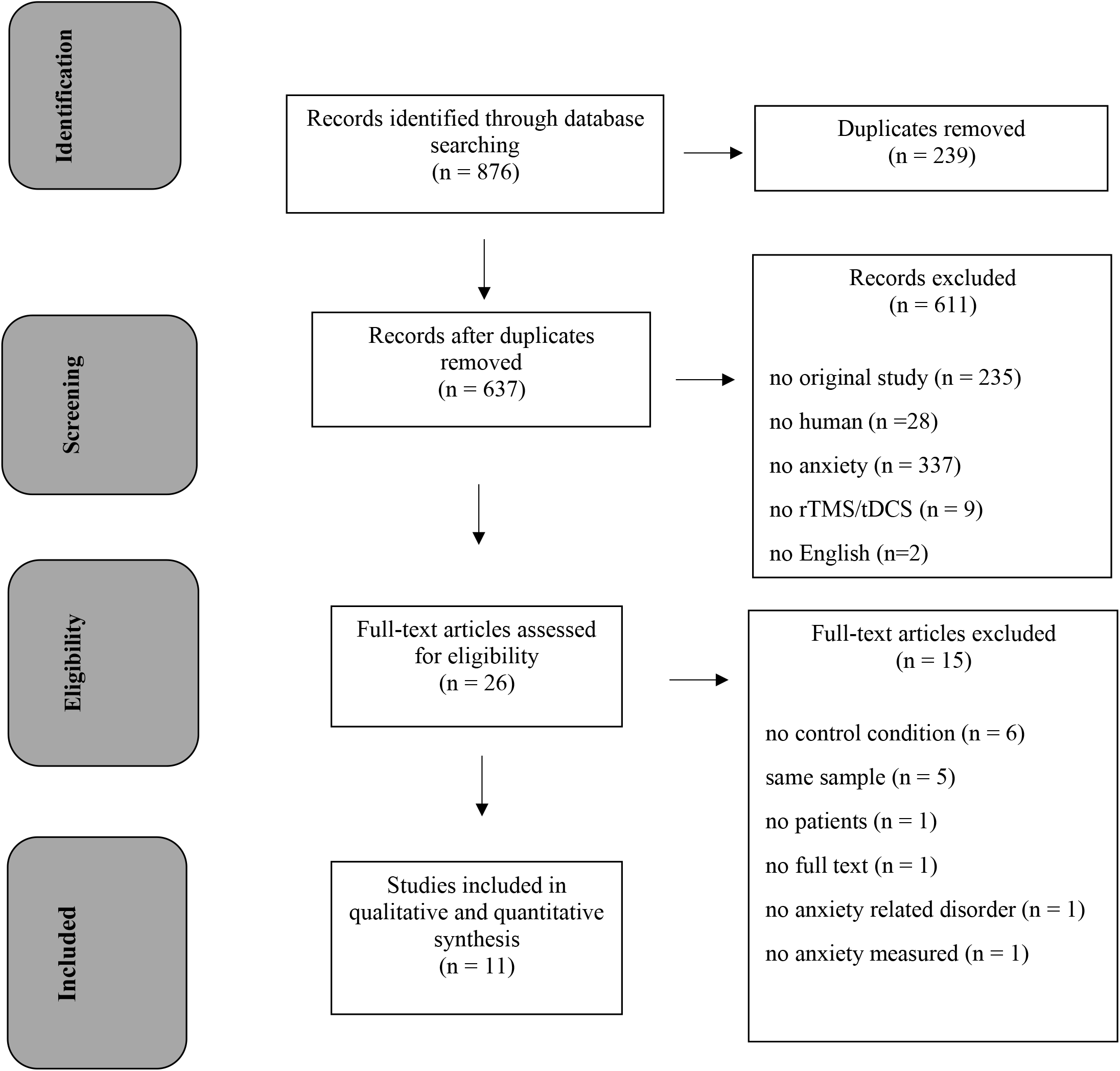
Flowchart of study selection process.

### Quality Assessment

Results of the quality assessment are reported in Table 3. We calculated the percentage of the high-risk judgments to obtain a quality score for each study. The average quality of the studies included in the present review was high to intermediate (range 0-42.86%), with the random sequence generation as the primary source of methodological bias, followed by the blinding mode. Indeed, most of the studies did not describe the randomization procedures, and two studies employed a single-blind design ^91, 117^. Finally, three studies ^90, 91, 118^ reported confusing information about the number of patients excluded from the final sample analyzed.

**Table 3.**
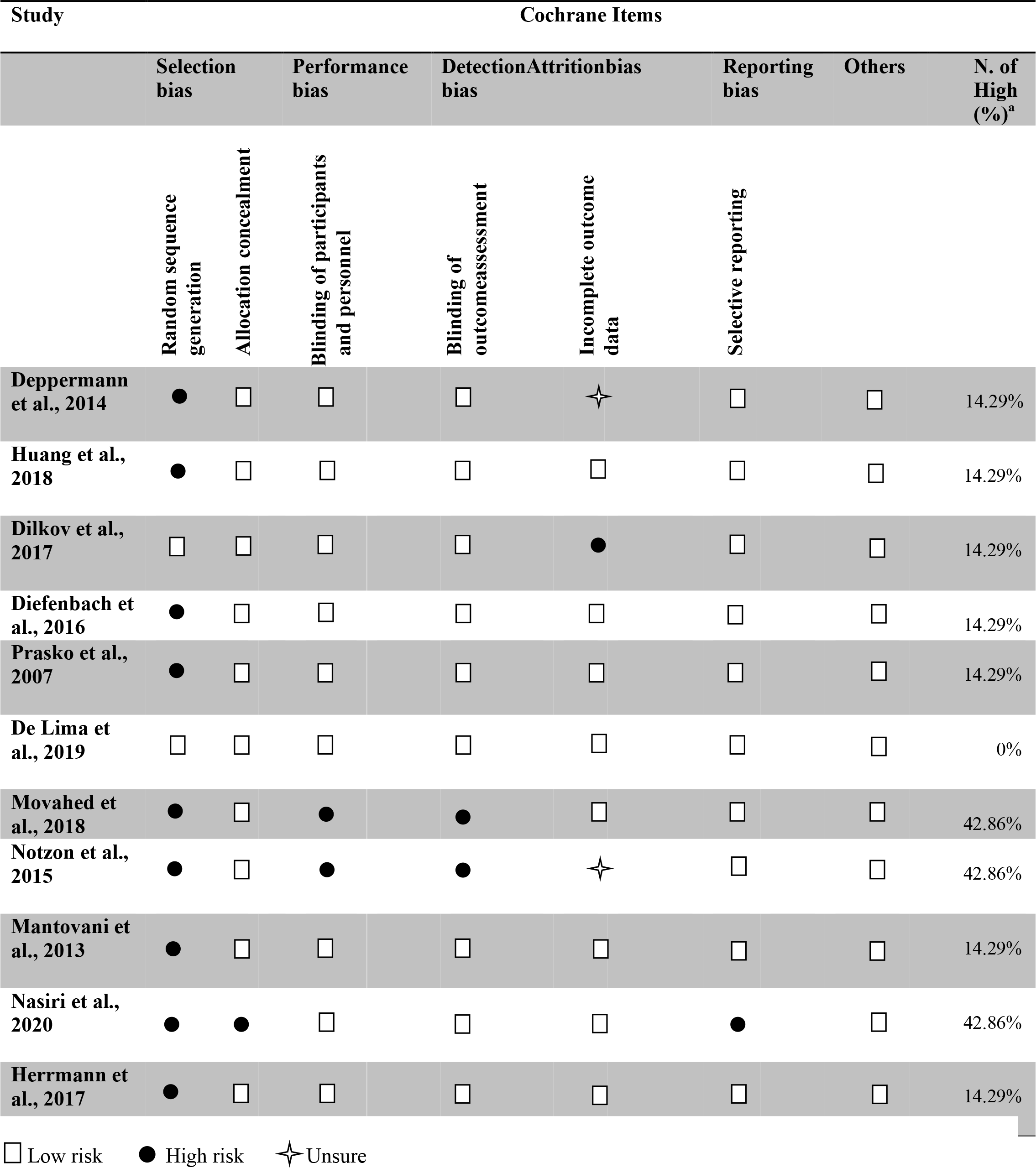
Risk of bias evaluation of papers included in the meta-analysis.

Reporting bias was evaluated considering the details reported in the full texts, except for three studies ^89, 119, 120^ whose registered protocol was available to check the completeness and the consistency of the findings. Among these, Nasiri and colleagues ^120^ did not report analyses and results of some of the pre-registered outcome variables, being part of a larger project. Concerning the allocation concealment, only Nasiri et al. ^120^ obtained a high-risk judgment as they used cognitive treatment and not sham stimulation as a control condition.

### Participants’ characteristics and inclusion/exclusion criteria

Eleven studies were included in the meta-analysis, involving 154 participants assigned to the experimental condition with real stimulation and 164 assigned to the control groups. Concerning demographic variables, participants’ age ranged between 18 and 65 years; when reported (10 out of 11 studies), the mean age of participants was 36.4 years for real stimulation conditions (SD± 6.6) and 36.8 (SD± 7.2) for control groups. In most studies, the number of females was greater than the number of males, and secondary school was the most common education level. Specific participants’ features of the included studies are reported in Table 4.

**Table 4.** Summarizes participants’ characteristics of the included studies. PD =panic disorder; GAD = generalized anxiety disorder; SP = specific phobia; MDD =major depressive disorder.

|  | Real stimulation |  |  |  | Control condition |  |  |  |  | Recruitment |
| --- | --- | --- | --- | --- | --- | --- | --- | --- | --- | --- |
| Author | Age | Males | Females | Education | Age | Males | Females | Education | Diagnosis |  |
| Deppermann et al., 2014 | Mean = 37.6 (range 19–63) | 9<sup>a</sup> | 13<sup>a</sup> | Mean = 12.1 (SD = 1.7) | 36.3 (range 22–56) | 8<sup>a</sup> | 14<sup>a</sup> | Mean = 12.4 (SD = 2.0) | PD ± agoraphobia | Outpatient clinics; advertisements; internet; information material sent to local physicians |
| Huang et al., 2018 | Mean = 44.94 (SD = 11.64) | 9 | 9 | n.r. | Mean 45.22 (SD = 10.85) | 9 | 9 | n.r. | GAD + insomnia | Neurology outpatient's clinic |
| Dilkov et al., 2017 | Mean = 34 (SD = ± 7) | 9 | 6 | n.r. | Mean = 38 (SD = ± 10) | 11 | 11 | n.r. | GAD | Two mood disorder centers: Canada and Bulgaria |
| Diefenbach et al., 2016 | Mean = 44.00 (SD = 11.95) | 1 | 8 | 12 (high school diploma) | Mean = 44.58 (SD = 14.75) | 3 | 7 | 12 (high school diploma) | GAD | Outpatient clinic; advertisements; internet; community flyers; physician referral; media coverage |
| Prasko et al., 2007 | Mean = 33.7 (SD = ± 9.2) | 1 | 6 | 5: 1: 1 (basic :secondary : university) | Mean = 33.8 (SD = ± 12.2) | 3 | 5 | 1 : 6 : 1 (basic : secondary : university) | PD | n.r. |
| De Lima et al., 2019 | Mean = 32.07 (SD = ± 6.5) | 5 | 10 | 2 (Elementary); 9 (Secondary); 4 (University) | Mean = 29 (SD = ± 5.05) | 6 | 9 | 2 (Elementary); 7 (Secondary); 6 (University) | GAD | Two outpatient clinics. |
| Movahed et al., 2016 | n.r. | n.r. | n.r. | n.r. | n.r. | n.r. | n.r. | n.r. | GAD | n.r. |
| Notzon et al., 2015 | Mean = 25.85 (SD = 7.65) | 20<sup>b</sup> |  | Mean = 11.30 (SD = 3.91) | 27.02 (9.23) | 20<sup>b</sup> |  | Mean = 11.34 (SD = 3.51) | SP | Local advertisement |
| Mantovani et al., 2013 | Mean = 40.2 (SD = 710) | 4<sup>c</sup> | 8<sup>c</sup> | n.r. | Mean = 39.87 (SD = 13.3) | 8<sup>c</sup> | 5<sup>c</sup> | n.r. | PD + MDD | n.r. |
| Nasiri et al., 2020 | Mean = 20.23 (SD = 2.89) | 3 | 10 | n.r. | Mean = 21.53 (SD = 3.56) | 4 | 11 | n.r. | GAD + MDD | University announcements |
| Herrmann et al., 2017 | Mean = 43.2 (SD = 12.6) | 7 | 13 | n.r. | Mean = 46.6 (SD = 13.7) | 6 | 13 | n.r. | SP | Advertisements in local newspapers |

<sup>a</sup>The number of males and females is based on the original number of participants included in the study reported in Deppermann et al., 2017. A total of 3 participants did not complete the study (2 for the real and 1 for the sham stimulation), but their gender is not reported by the authors.
<sup>b</sup>The participants' gender in the real and sham condition is not specified, therefore we reported the total number of patients included in the authors' dataset.
<sup>c</sup>The number of males and females is based on the original number of participants included in the study. A total of 4 participants did not complete the study (1 for the real and 3 for the sham stimulation).

The studies differed regarding the number of stimulation sessions (ranging from 1 to 25 sessions), the intervention technique (rTMS, tDCS, intermittent theta-burst stimulation, or iTBS) (see the “stimulation intervention” paragraph), the presence or absence of concomitant treatments (pharmacological or psychological interventions, see the “associated therapies” paragraph), and the patients’ diagnosis.

Concerning inclusion and exclusion criteria, they changed across studies. Participants were typically included if they were in a certain age range, had a specific diagnosis (according to DSM) and reached a certain questionnaire score. Conversely, exclusion criteria typically concerned prior psychiatric history - except for the actual disorder - and suicidality (see Table 7 for details).

The following paragraph will report the details of the samples included.

### Patients’ diagnosis

Studies were included if participants received a primary diagnosis of anxiety disease, which could be in comorbidity with depression. Studies in which anxiety was secondary to other conditions (e.g., organic/neurological condition, substance use, etc.) were excluded. Concerning the eleven included studies, participants had a diagnosis of generalized anxiety disorder in 6 out of 11 studies ^89, 90, 117, 119–121^ - in 1 out of 6 combined with insomnia ^121^ and in another one with major depression disorder ^120^. In 3 out of studies participants had panic disorder with or without agoraphobia ^118, 122, 123^ - in 1 out of 3 major depression disorder was in comorbidity ^122^. The last two papers included participants with specific phobia, namely spider phobia ^91^ and acrophobia ^124^.

### Associated therapies

Concerning the associated psychological interventions, 4 out of 11 studies ^91, 118, 120, 124^ provided psychological intervention as part of the treatment. Specifically, in Deppermann et al. ^118^, participants took part in three group sessions of psychoeducation concerning the panic disorder, which occurred separately from the stimulation sessions. Nasiri et al. ^120^ added the NIBS treatment to the last two weeks of a 12 unified protocol ^125^ (UP^3^) weekly sessions, without indicating whether the simulation was time locked (e.g., during or immediately before) to the psychological intervention. Notzon et al. ^91^ and Herrmann et al. ^124^ applied the stimulation before virtual reality exposure, even though the interventions were occurred in a single and double sessions, respectively. In 2 out of 11 studies ^90, 122^, individual or supportive psychotherapy was allowed during NIBS sessions, while in 4 out of 11 studies ^89, 117, 119, 121^, psychological interventions were not permitted during NIBS treatment. One study^123^ did not report information about this aspect.

Concerning concurrent pharmacotherapy treatment, 4 out of 11 studies ^91, 117, 124^ did not allow medication intake during the NIBS treatment, while the other 7 ^89, 90, 118, 119, 121–123^ reported that stable medication treatment was accepted. Medication stability was differently defined across the studies, ranging from 4 weeks before treatment onset ^122^ to the three months before ^119, 121^ (see Table 5 for details).

**Table 5.**
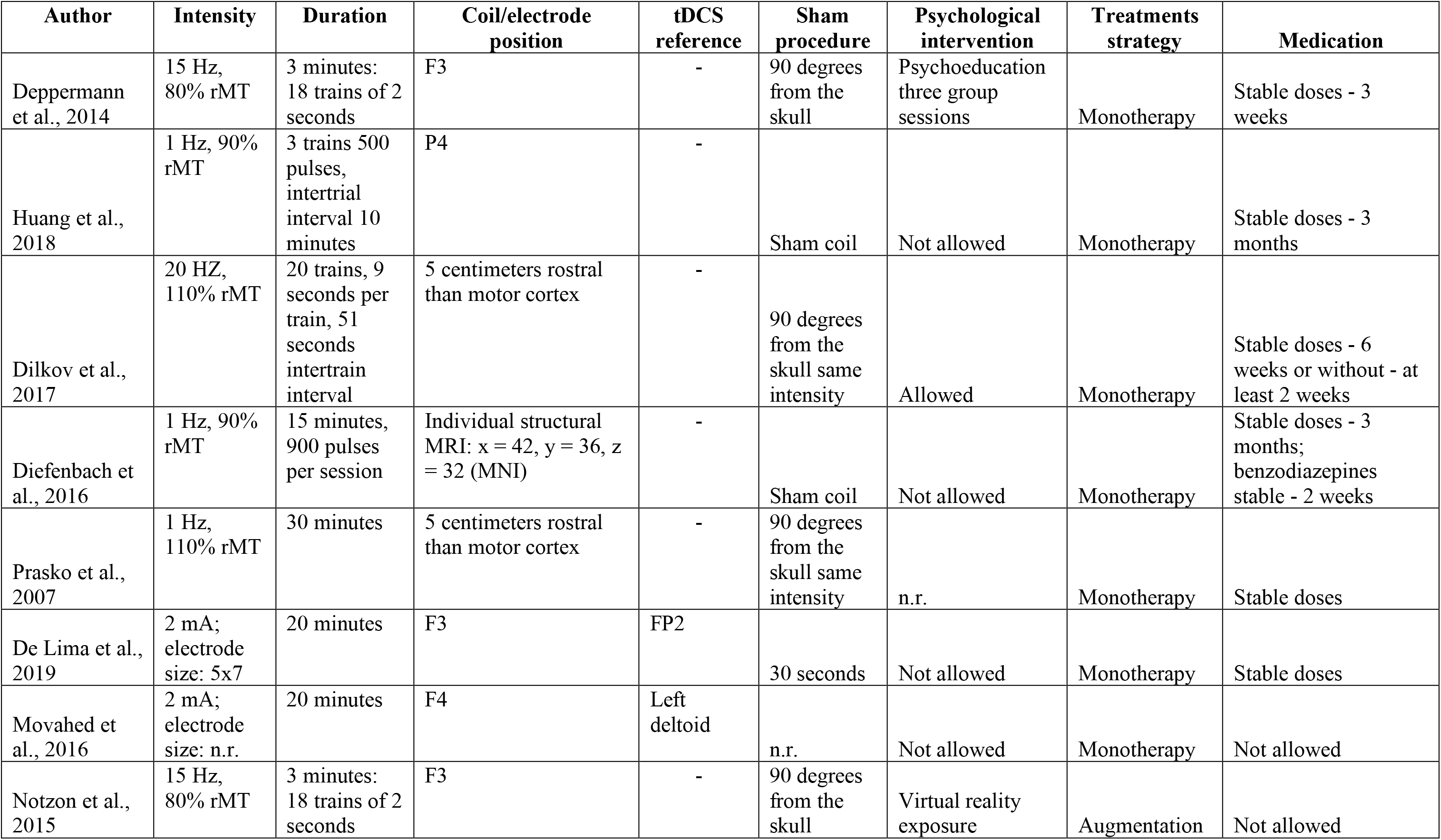
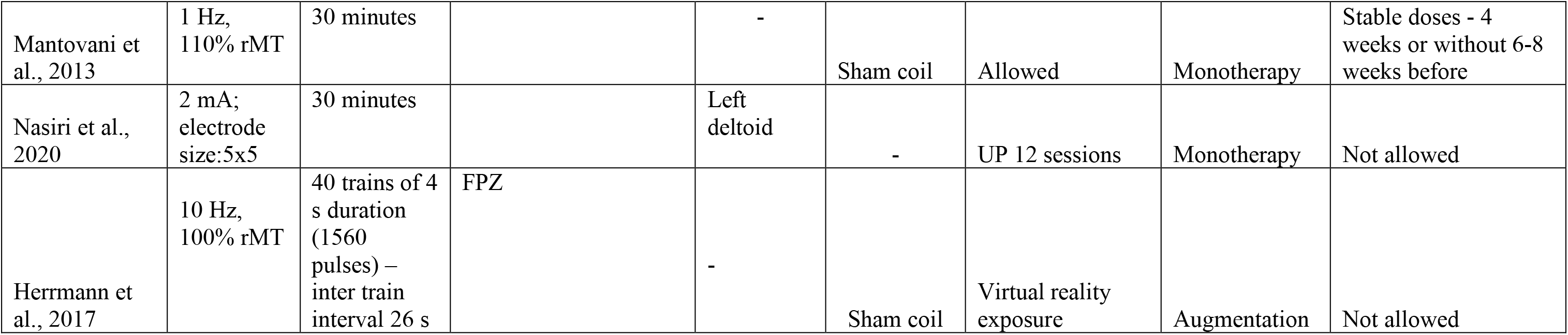
Summarizes stimulation protocols details, treatment strategy and associated therapies. Hz = hertz; rMT = repetitive motor threshold; mA = milliAmpere; F3 = 10-20 EEG position corresponding to the left dorsolateral prefrontal cortex; F4 = 10-20 EEG position corresponding to the right dorsolateral prefrontal cortex; P4 = 10-20 EEG position corresponding to the right posterior parietal cortex; FP2 = 10-20 EEG position corresponding to the supraorbital region; FPZ = 10-20 EEG position corresponding to the ventromedial prefrontal cortex; UP = Unified Protocol for Transdiagnostic Treatment of Emotional Disorders; n.r. = not reported.

### Stimulation protocols

Of the final eleven studies, 6 used an rTMS protocol ^90, 119, 121–124^, three a tDCS ^89, 117, 120^, and two an iTBS protocol ^91, 118^. RTMS was applied at a frequency of 1 Hz in 4 out of 6 studies ^119, 121–123^, at 20 Hz in 1 study ^90^ and 10 Hz in another one ^124^. The target region for rTMS intervention was the right DLPFC in 4 out of 6 articles, the right posterior parietal cortex (PPC) for 1 study ^121^ and the ventromedial prefrontal cortex (vmPFC) for the latter one ^124^. The two iTBS protocols were applied over the left DLPFC. Concerning the three tDCS studies, stimulation was delivered with the cathodal polarity over the right DLPFC in 2 out of 3 studies ^117, 120^ and with anodal polarity over the left DLPFC in the remaining one ^89^. Overall, inhibitory protocols (cathodal tDCS, 1 Hz rTMS) were applied over the right DLPFC in 5 out of 6 studies, while only one targeted the right PPC. Facilitatory protocols (iTBS, anodal tDCS, and 20 Hz rTMS) were delivered over the left DLPFC in 3 out of 5 studies, over the right DLPFC in 1 study (see Figure 1 for a graphical representation of targeted regions) and over the vmPFC in one study. Stimulation intensity range in TMS studies was between the 80% and the 110% of the individual rest motor threshold (RMT). Magnetic pulses were delivered with a figure of eight shaped coils except for the study by Herrmann and colleagues ^124^ in which a round coil was used. tDCS protocols were administered at 2mA intensity in the three protocols, with unipolar montages and intracephalic reference in 1 out of 3 studies ^89^ and a deltoid reference in 2 out of 3 protocols ^117, 120^. Stimulation duration ranged from 10 to 30 minutes. See Tables 5 and 6 for details.

**Figure 1.**
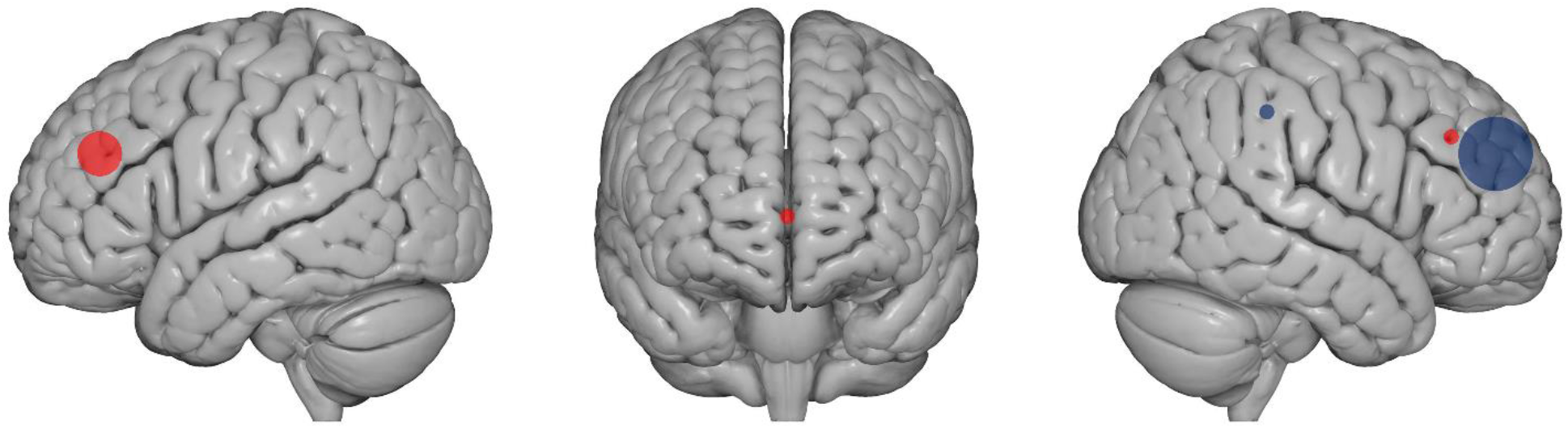
summarizes the type of stimulation and target regions of included studies. Red dots indicate excitatory stimulation protocols (i.e., anodal tDCS, iTBS and high frequency rTMS), while blue dots indicate inhibitory stimulation (i.e., cathodal tDCS and low frequency rTMS). Dots’ size corresponds to the number of studies applying excitatory or inhibitory protocol over a specific region: five studies applied inhibitory protocols over the right DLPFC, three delivered excitatory protocols to the left DLPFC, one excitatory stimulation over the right DLPFC, one inhibitory stimulation over the right PPC, one excitatory stimulation over the vmPFC. Brain images were obtained from www.nitrc.org.

**Table 6.** Summarizes stimulation protocol, statistical analyses, main results, additional groups and measures. fNIRS = functional near-infrared spectroscopy; rTMS = repetitive transcranial magnetic stimulation; ANOVA = Analysis of Variance; MANCOVA = Multivariate analysis of covariance; CAQ = Cardiac Anxiety Questionnaire; CGI = Clinical Global Impression Scale; DASS-DEP = Depression-Anxiety Scales-Depression Subscale; fMRI = functional magnetic resonance imaging; BAI = Beck Anxiety Inventory; ANAS = Positive and Negative Affect Schedule; FSQ = Fear of Spiders Questionnaire; ASI = Anxiety Sensitivity Index; DS = Disgust Scale; SUDS = Subjective Units of Discomfort Scale; IPQ = Igroup Presence Questionnaire; HR = heart rate; SCL = skinconductance level; BDI =Beck Depression Inventory; PGI = Patient Global Impression; SASS = Self-reported Social Adaptation Scale; PDSS (-SR) = Panic Disorder Severity Scale (self-report); ZUNG - SAS= Zung-Self Administered Scale;IUS = Intolerance of Uncertainty Scale; PSWQ = Penn State Worry Questionnaire; AQ = Acrophobia questionnaire; BAT = Behavioral Avoidance Test; n.r. = not reported.

| **Author** | **Protocol** | **Follow up** | **Statistical analysis** | **Reported results** | **Additional groups** | **Additional pre-post measures** |
| --- | --- | --- | --- | --- | --- | --- |
| Deppermann et al., 2014 | 5 daily sessions - 3 weeks | n.r. | ANOVA repeated measures | No differences in real vs. sham rTMS. Both groups showed anxiety symptoms improvement in post iTBS vs. baseline measurements. | Healthy controls - only for fNIRS | Physiological: CAQ<br>Brain activation: fNIRS<br>Cognitive: verbal fluency |
| Huang et al., 2018 | 10 consecutive days | 2: 2 weeks, 1 month | ANOVA repeated measures | Anxiety, insomnia and depressive symptoms improvement in real vs. sham rTMS at the end of the treatment and at the two follow-ups. | Not present | n.r. |
| Dilkov et al., 2017 | 6 weeks - 5 sessions a week for the first 4 weeks; during the 5th week, sessions were reduced to 3 times/week and again to twice a week during the 6th week. | 2: 2, 6 weeks after the end of the treatment | ANOVA repeated measures | Anxiety and depressive symptoms improvement in the real vs. sham condition at the end and at the two follow-ups. | Not present | Global evaluation: CGI |
| Diefenbach et al., 2016 | 5 daily sessions - 6 weeks | 2: 3 months, 6 months (only a subset not included in statistical analysis) | ANOVA repeated measures; planned contrasts | Anxiety symptoms improvements in post vs. pre measurements in real and sham rTMS, that persisted at the 3 months follow-up only in the real rTMS.<br>Worry and depressive symptoms improved only in the real rTMS at the end of the treatment and at the 3-months follow-up. | Not present | Anxiety/mood: DASS-DEP<br>Brain activation: fMRI during gambling task |
|  |  |  |  | Brain activation increased after real rTMS and tended to decrease after sham rTMS. |  |  |
| Prasko et al., 2007 | 5 daily sessions - 2 weeks | 1: 2 weeks later | Non-parametric repeated measure analysis of variance. | Anxiety symptoms and psychopathology global scores improved after both real and sham rTMS. | Not present | Anxiety: BAI<br>Global evaluation: CGI |
| De Lima et al., 2019 | 5 consecutive days | 1: 1 week later | ANOVA repeated measures | Anxiety and depression symptoms did not differ between real and sham tDCS. Physical symptoms of stress reduced at the end and the follow-up in the real tDCS as compared to sham tDCS. | Not present | Anxiety: BAI<br>Global evaluation: PANAS |
| Movahed et al., 2016 | 4 weeks | 1: 2 months | ANOVA repeated measures | Worry, anxiety and depression scores reduced after cathodal tDCS and pharmacotherapy vs. sham tDCS. Pharmacotherapy was stronger than tDCS in reducing worry, while tDCS was stronger in reducing depression. Anxiety symptoms did not differ after cathodal tDCS compared to pharmacotherapy. | Pharmacotherapy | n.r. |
| Notzon et al., 2015 | Single session | n.r. | ANOVA repeated measures | itBS increased sympathetic activity during the spider scene in both phobic and healthy participants. | Healthy controls (real and sham) | Anxiety: FSQ; ASI<br>Global evaluation: IPQ; SUDS; DS<br>Physiological: HR; SCL<br>Brain activation: fNIRS |
| Mantovani et al., 2013 | 5 days a week - 4 weeks double blind + 4 weeks real* | 3: 1, 3, 6 months | ANOVA repeated measures; t-test | 4 weeks rTMS vs. sham: improvement in panic symptoms but not depression. | Not present | Anxiety: PDSS, PDSS-SR;<br>Mood: BDI; ZUNG – SAS |
|  |  |  |  | 8 weeks of rTMS vs. pre-treatment: improvement in panic and depressive symptoms, global assessment, and social adjustment. |  | Global evaluation: CGI; PGI; SASS |
| Nasiri et al., 2020 | 10 daily sessions -2 weeks | 1: 3 months later | MANCOVA | Worry, anxiety and anxiety sensitivity improved after UP + tDCS intervention as compared to UP alone at the end of the treatment and at follow-up. | waiting list | Anxiety: ASI; IUS; PSWQ |
| Herrmann et al., 2017 | 2 sessions | 1: 3 months later | ANOVA repeated measures; t-test | Two sessions of rTMS reduced anxiety and avoidance ratings as compared to the sham group. | Not present | Anxiety: AQ- avoidance subscale; BAT; |

\*We included in our analysis the data of the baseline and of the first 4 weeks of rTMS treatment.

**Table 7.** summarizes inclusion and exclusion criteria of the selected studies.

| Author | Participants inclusion criteria | Exclusion criteria |
| --- | --- | --- |
| Deppermann et al. 2014 | i) age 18-65;<br>ii) PD with or without agoraphobia according to DSM-IV-TR; | severe somatic disorders |
| Huang et al. 2018 | i) age 18-65;<br>ii) GAD primary diagnosis according to DSM IV;<br>iii) insomnia at least 3 months | i) prior history of other psychiatric diseases except GAD;<br>ii) concurrent psychotherapy or counseling |
| Dilkov et al. 2017 | i) age 18-65;<br>ii) GAD primary diagnosis according to DSM IV; | i) diagnosis of psychotic, bipolar I, MDD or substance/alcohol dependence in the 6 months before the study;<br>ii) severe axis II disorder;<br>iii) suicidal;<br>iv) severe or unstable medical conditions;<br>v) ECT treatment in the three previous months;<br>vi) TMS treatment in the 6 months before. |
| Diefenbach et al. 2016 | i) age higher than 18;<br>ii) GAD as principal or coprincipal disorder<br>iii) HRSA and HRSD cut off; | i) unstable medical/psychiatric condition (e.g. thyroid disease, suicidality);<br>ii) current PTSD;<br>iii) substance use disorder;<br>iv) lifetime bipolar, psychotic, developmental or obsessive–compulsive disorder;<br>v) concurrent psychotherapy |
| Prasko et al. 2007 | i) ICD-10 PD with/without agoraphobia;<br>ii) non-responders on SRIs (at least 6 weeks);<br>iii) age 18–45 years | i) MDD;<br>ii) suicidality;<br>iii) HAMD higher than 16;<br>iv) organic psychiatric disorder;<br>v) psychotic disorder in history;<br>vi) abuse of alcohol or other drugs;<br>vii) serious somatic disease;<br>viii) using non-prescribed medication |
| De Lima et al. 2019 | i) GAD diagnosis according to DSM 5;<br>ii) age between 20- 30 y.o. | psychotherapy or hospitalization indication from the psychiatrist at the beginning of the study; |
| Movahed et al. 2018 | i) GAD diagnosis according to DSM 5;<br>ii) age between 18- 55 y.o.;<br>iii) 5 points or higher in the 7-item GAD scale | i) previous mental illness;<br>ii) current physical illness;<br>iv) current psychological or pharmacological medication |
| Notzon et al. 2015 | i) age 18-65;<br>ii) spider phobia diagnosis according to DSM-IV-TR;<br>iii) at least 16 SPQ | i) severe somatic disorders;<br>ii) history of psychiatric disorders except for specific phobia;<br>iii) psychiatric or psychotropic medication |
| Mantovani et al. 2013 | i) age 18-65; | i) suicidal risk; |
|  | ii) PD and MDD primary diagnosis of DSM-IV-TR;<br>iii) current episode duration of at least a month;<br>iv) having residual panic attack and MDD symptoms despite medication;<br>v) stable medication for 4 weeks;<br>vi) stable psychotherapy (3 months). | ii) history of bipolar or psychotic disorders, substance, or dependence abuse within the previous year |
| Nasiri et al. 2020 | i) GAD primary diagnosis according to DSM-V;<br>ii) comorbid MDD diagnosis according to DSM-V;<br>iii) no medication use;<br>iv) age 18-40;<br>v) speak Persian fluently;<br>vi) ability to participate in all assessment and treatment sessions. | i) need for immediate medical/therapeutic interventions;<br>ii) receiving no more than 8 sessions of CBT-based interventions within the last 5 years;<br>iii) having psychiatric disorders/substance abuse;<br>iv) current diagnosis of mental disorders;<br>v) opposition to collaboration at any time of research;<br>vi) suicidality;<br>vii) history of receiving other psychological treatments. |
| Herrmann et al. 2017 | i) specific phobia (acrophobia) diagnosis according to DSM-IV;<br>ii) subjective motivation to do something about their fear (at least “3” on a scale from 0 to 10 – extreme motivation)<br>iii) motion sickness with 3D movies below 4 (on a scale 0 -10) | i) heights treatment within the previous 6 months;<br>ii) concurrent involvement in psycho- or pharmacotherapy. |

### Control condition

The presence of a control condition was established as an inclusion criterion in our meta-analysis. For 10 out of 11 studies, it consisted of a sham condition. In contrast, in one study ^120^, the control group did not receive a sham stimulation but took part in the UP treatment. For rTMS studies, sham stimulation was induced by varying the coil inclination at 90° with respect to the stimulation site in 4 out of 8 studies ^90, 91, 118, 123^. In contrast, in 4 out of 8 studies ^119, 121, 124, 126^, experimenters used a sham coil, which had the same appearance and produced the same noise as the real one. Considering the three tDCS studies, 1 applied the typical sham-tDCS protocol ^89^, namely the stimulation was on for the first 30 seconds, thus inducing the same skin sensation as compared to the real one ^127^; Movahed and colleagues ^117^ did not report the sham protocol parameters, and Nasiri et al. ^120^ did not have a sham condition but included a control group, in which participants did not receive sham stimulation, but only take part to the cognitive treatment.

Considering the blinding procedure, 9 out of 11 protocols were double-blind, with both experimenters and participants naïfs to participants’ assigned condition. In contrast, 2 out of 11 studies ^91, 117^ used a single-blind design, in which only participants were naïfs to the stimulation group. In 4 out of 11 studies, participants’ blinding was checked with specific questionnaires ^91, 118, 122, 124^.

### Outcome measures

As previously reported, we chose three outcome measure, namely i) a specific anxiety measure centered on the specific disorder investigated in each study, which was reported in 10 out of 11 studies (all but ^90^); ii) a general anxiety measure, investigating general anxiety symptoms, reported in 9 out of 11 studies (all but ^91, 124^); iii) a measure of depression, which was included in 7 out of 11 studies (depression questionnaires were not included in ^91, 118, 123, 124^).

The specific anxiety outcome measure included scores from a heterogeneous pool of clinical validated questionnaires, depending on the specific disease. For the panic disorder, 2 out of 3 studies ^122, 123^ administered the Panic Disorder Severity Scale ^85^ (PDSS), and the other one ^118^ the Panic and Agoraphobia Scale ^128^ (PAS). For the specific phobia studies, the German version of the Spider Phobia Questionnaire ^129, 130^ (SPQ) was included by Notzon and colleagues ^91^, while Herrmann et al.^124^ included the German translation of the Acrophobia Questionnaire ^131^ (AQ) – anxiety subscale. Concerning generalized anxiety disorder, the Penn State Worry Questionnaire ^132^ (PSWQ) was included for 2 out of 6 studies ^117, 119^, the Generalized Anxiety Disorder Questionnaire ^133^ (GAD-Q-IV) was included for one study ^120^, the Lipp Inventory of Stress Symptoms for Adults ^134^ (ISSL) for another one ^89^, and the Pittsburgh Sleep Quality Index ^135^ (PSQI), investigating insomnia symptoms, for the fifth ^121^. The sixth GAD study ^90^ did not include a disorder-specific questionnaire; therefore, it was not included in the specific anxiety disorders analysis (see Results section).

Concerning the general anxiety measure, for 8 out of 9 studies we included the Hamilton anxiety rating scale, HAM-A ^83^, which is a 14-item clinical interview targeting somatic and psychic anxiety symptoms. For 1 out of 9 studies ^120^, we included the Beck Anxiety Inventory, BAI ^136^, a 21-items self-report questionnaire focusing on the somatic symptoms of anxiety occurring over the past week. Notzon and colleagues ^91^ study did not include a general anxiety measure; therefore, it was not included in the analysis of general indexes of anxiety (see Results section).

Considering depression outcome measure, for 5 out of 7 studies ^90, 117, 119, 121, 122^ we included the Hamilton Depression Rating Scale, HAM-D ^86^ a 21-items (but only the first 17 concurred to the total score) clinical interview targeting somatic and neurovegetative aspects of depression, and for 2 out of 7 ^89, 120^ the Beck Depression Inventory (BDI) ^87^ a 21-items self-report questionnaire investigating disorder’s cognitive and affective dimensions (for a comparison between HAM-D and BDI, see ^137^). When both the clinical and the self-report measures of general anxiety or depression were reported only the clinician-administered version was considered.

### Meta-analyses results

#### Anxiety specific disorders

Ten out of eleven studies reported scores at specific anxiety disorders scales (see Table 1). A meta-analysis was run on these studies to compute the global effect of NIBS on the reduction of anxiety specific symptoms compared to a sham intervention. The random-effects model showed a significant medium impact of non-invasive brain stimulation on patients’ improvement compared to pre-post sham scores (overall SMD of -.49; 95% CI = [-.83, -.14], p=.006; see Table 8 for the complete results and Fig. 2 for the forest plot).

**Figure 2.**
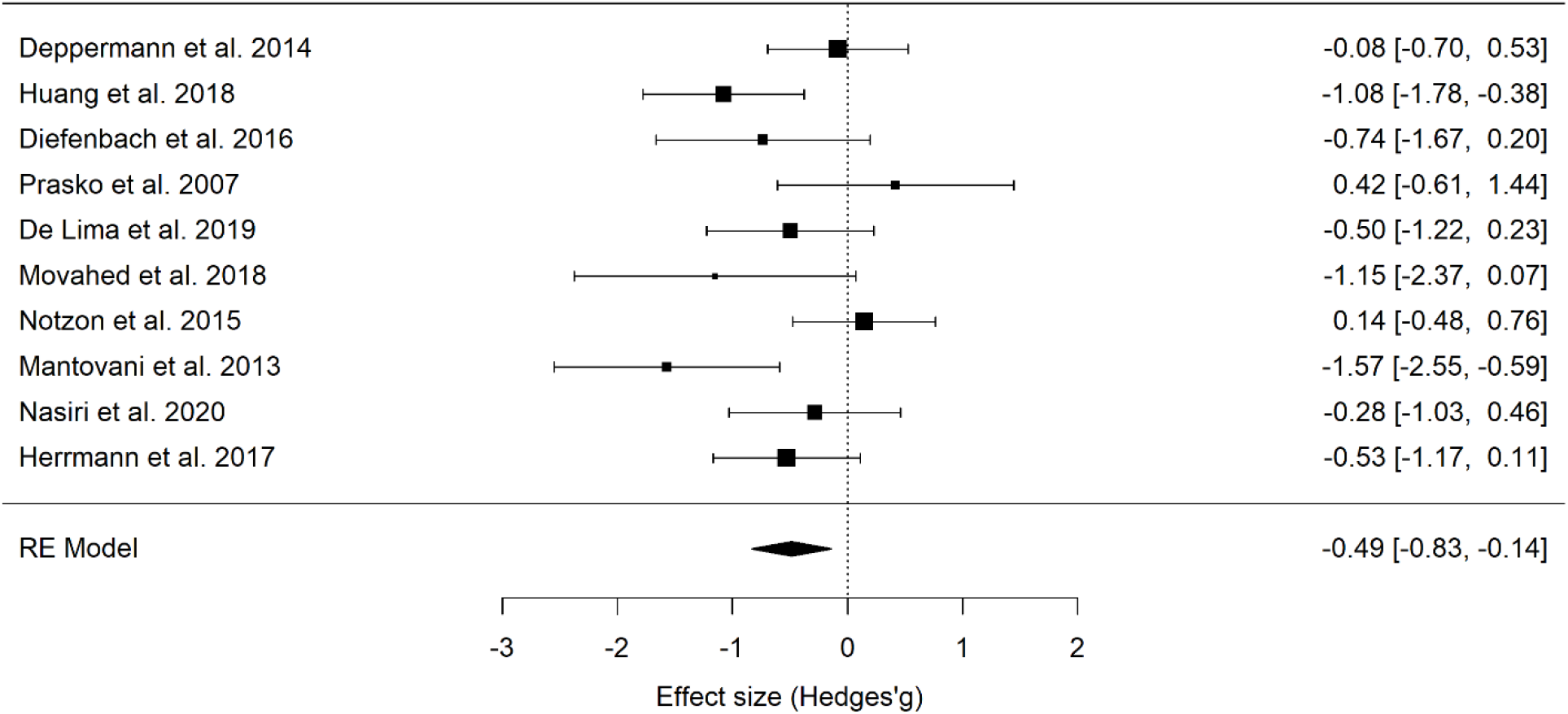
Forest plot of the effect size of NIBS on continuous specific anxiety questionnaire scores.

**Table 8.**
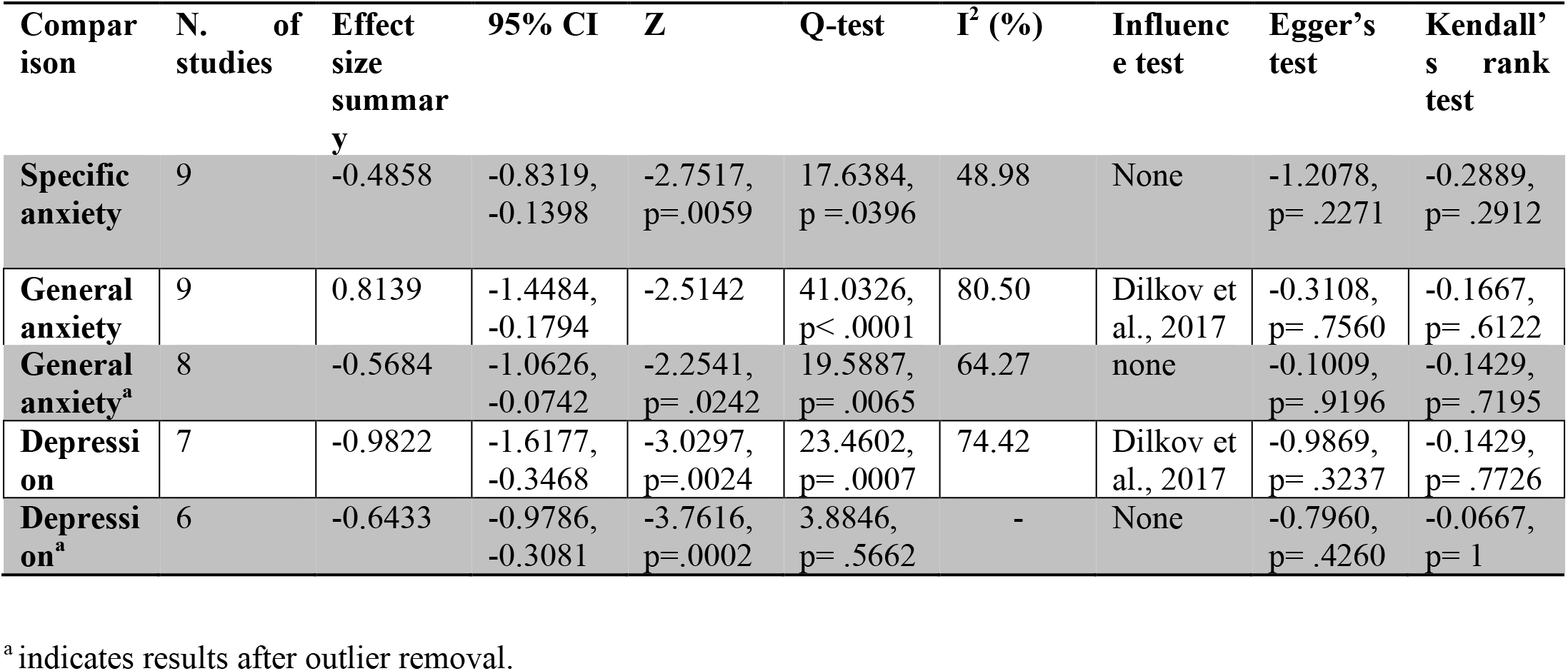
summarizes results of the three meta-analyses.

| Comparison | N. of studies | Effect size summary | 95% CI | Z | Q-test | I<sup>2</sup> (%) | Influence test | Egger's test | Kendall's rank test |
| --- | --- | --- | --- | --- | --- | --- | --- | --- | --- |
| **Specific anxiety** | 9 | -0.4858 | -0.8319, -0.1398 | -2.7517, p=.0059 | 17.6384, p=.0396 | 48.98 | None | -1.2078, p=.2271 | -0.2889, p=.2912 |
| **General anxiety** | 9 | 0.8139 | -1.4484, -0.1794 | -2.5142 | 41.0326, p<.0001 | 80.50 | Dilkov et al., 2017 | -0.3108, p=.7560 | -0.1667, p=.6122 |
| **General anxiety<sup>a</sup>** | 8 | -0.5684 | -1.0626, -0.0742 | -2.2541, p=.0242 | 19.5887, p=.0065 | 64.27 | none | -0.1009, p=.9196 | -0.1429, p=.7195 |
| **Depression** | 7 | -0.9822 | -1.6177, -0.3468 | -3.0297, p=.0024 | 23.4602, p=.0007 | 74.42 | Dilkov et al., 2017 | -0.9869, p=.3237 | -0.1429, p=.7726 |
| **Depression<sup>a</sup>** | 6 | -0.6433 | -0.9786, -0.3081 | -3.7616, p=.0002 | 3.8846, p=.5662 | - | None | -0.7960, p=.4260 | -0.0667, p=1 |

<sup>a</sup> indicates results after outlier removal.

Q-statistic and I^2^ suggest a high heterogeneity between studies (Tab. 8), and this may be due to the differences in methodological aspects across studies. The inclusion of moderators (stimulation sessions number, specific NIBS technique used, protocol type) was not significant (all ps> .128, see Table 9 for the moderators’ statistical results).

**Table 9.** shows the results of the moderation analysis for specific and general anxiety scores and depression scores. The applied technique (iTBS, rTMS, tDCS), target region (left vs. right DLPFC), protocol type (excitatory vs. inhibitory) moderators are categorical variables, while session number is a numerical variable. Only for the depression outcome measure, we computed whether the presence of comorbid depression influences the outcome of the scores. *Note*: *smd* = effect size. LL = lower limit of the 95% CI; UL = upper limit of the 95% CI; *z* = *z*-score associated with the *smd* value; *p* = *p*-value associated with the *z*-score in the same row; *Q* = result of the *Q*-test for moderation; *df* = degrees of freedom of the *Q*-test for moderation; *p* = *p*-value of the *Q*-test for moderation. ^†^*p* < .10.

| *Moderator* | *SMD* | *LL* | *UL* | *z* | *p* | *q* | *df* |
| --- | --- | --- | --- | --- | --- | --- | --- |
| ***Specific anxiety measure*** |  |  |  |  |  |  |  |
| *Session number* | -0.0414 | -0.1038 | 0.0209 | -1.3019 | .1929 | 1.6950 | 1 |
| *Technique* | -0.2827 | -0.7443 | 0.1788 | -1.2006 | .2299 | 1.4415 | 1 |
| *Target region* | -0.4963 | -1.2778 | 0.2852 | -1.2447 | .2132 | 1.5493 | 1 |
| *Protocol type* | -0.4965 | -1.1366 | 0.1435 | -1.5205 | .1284 | 2.3118 | 1 |
| ***General anxiety measure*** |  |  |  |  |  |  |  |
| *Session number* | -0.0723 | -0.1811 | 0.0364 | -1.3039 | .1923 | 1.7001 | 1 |
| *Technique* | -0.1830 | -1.2449 | 0.8790 | -0.3377 | .7356 | 0.1140 | 1 |
| *Target region* | -0.8212 | -2.2992 | 0.6568 | -1.0890 | .2762 | 1.1858 | 1 |
| *Protocol type* | 0.2243 | -1.2106 | 1.6592 | 0.3064 | .7593 | 0.0939 | 1 |
| ***Depression measure*** |  |  |  |  |  |  |  |
| *Session number* | -0.0777 | -0.1634 | 0.0080 | -1.7760 | .0757<sup>†</sup> | 3.1542 | 1 |
| *Technique* | 0.5794 | -0.7260 | 1.8847 | 0.8699 | .3844 | 0.7567 | 1 |
| *Target region* | -0.6709 | -2.9417 | 1.5998 | -0.5791 | .5625 | 0.3354 | 1 |
| *Protocol type* | 0.8540 | -0.5639 | 2.2718 | 1.1805 | .2378 | 1.3935 | 1 |
| *Comorbidity* | 0.9563 | -0.3677 | 2.2803 | 1.4157 | .1569 | 2.0042 | 1 |

Baujat plot inspection ^138^ (Figure 3) suggests that study 8 ^122^ greatly contributed to meta-analysis heterogeneity. Nevertheless, testing for a possible outlier influence of the included studies in the results ^94^ showed no study significantly differed from the rest of the data (Table 8). Concerning publication bias, the funnel plot (Fig. 4) showed no asymmetry according to both Egger’s regression test (z = −1.21; p=.227) and the Rank correlation analysis (Kendall’s tau = −0.29; p = 0.291).

**Figure 3.**
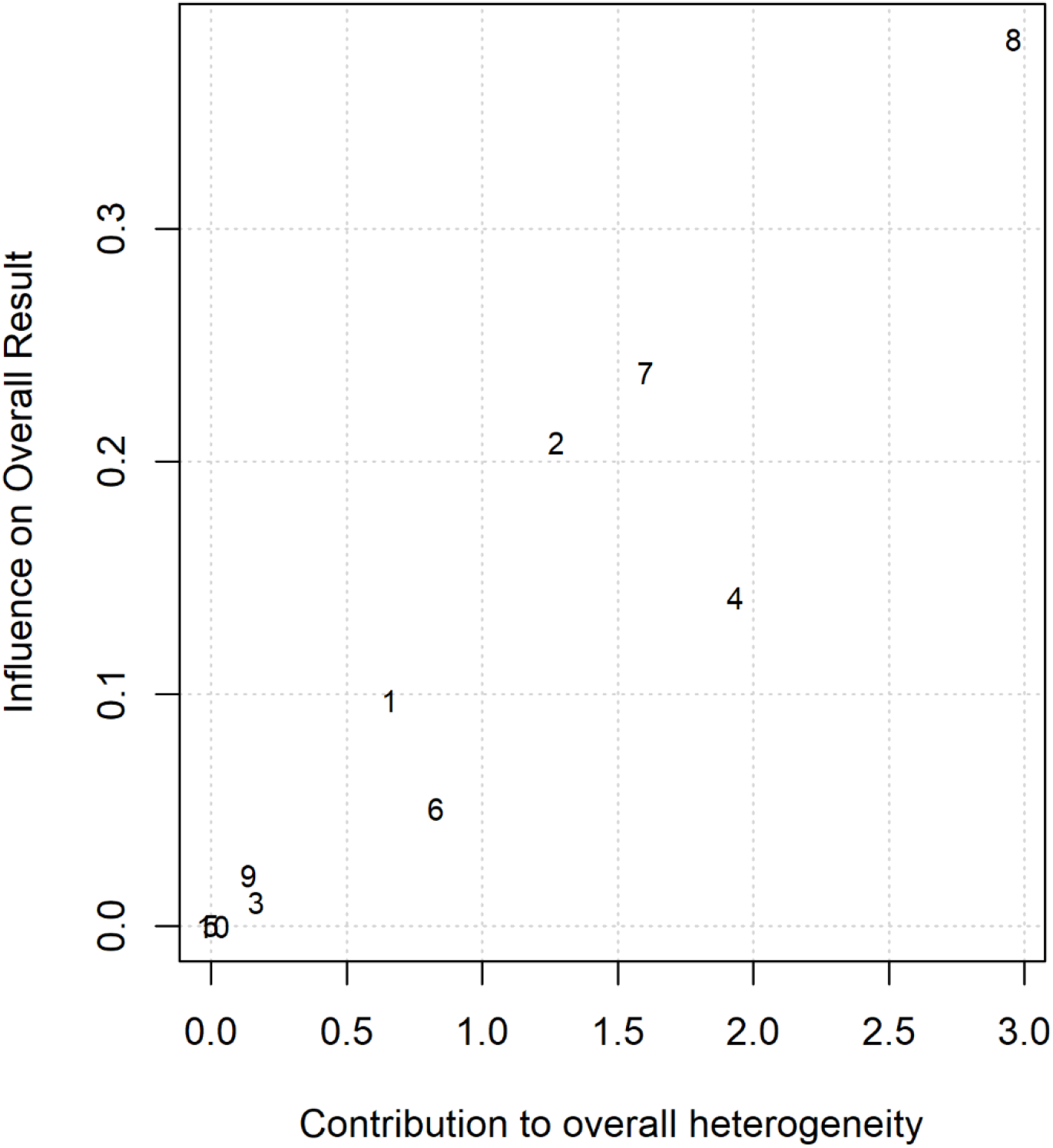
Baujat plot of studies distribution in terms of heterogeneity. At a visual inspection, study 8 (Mantovani et al., 2013) seems to contribute to the statistical heterogeneity in the analyzed studies.

**Figure 4.**
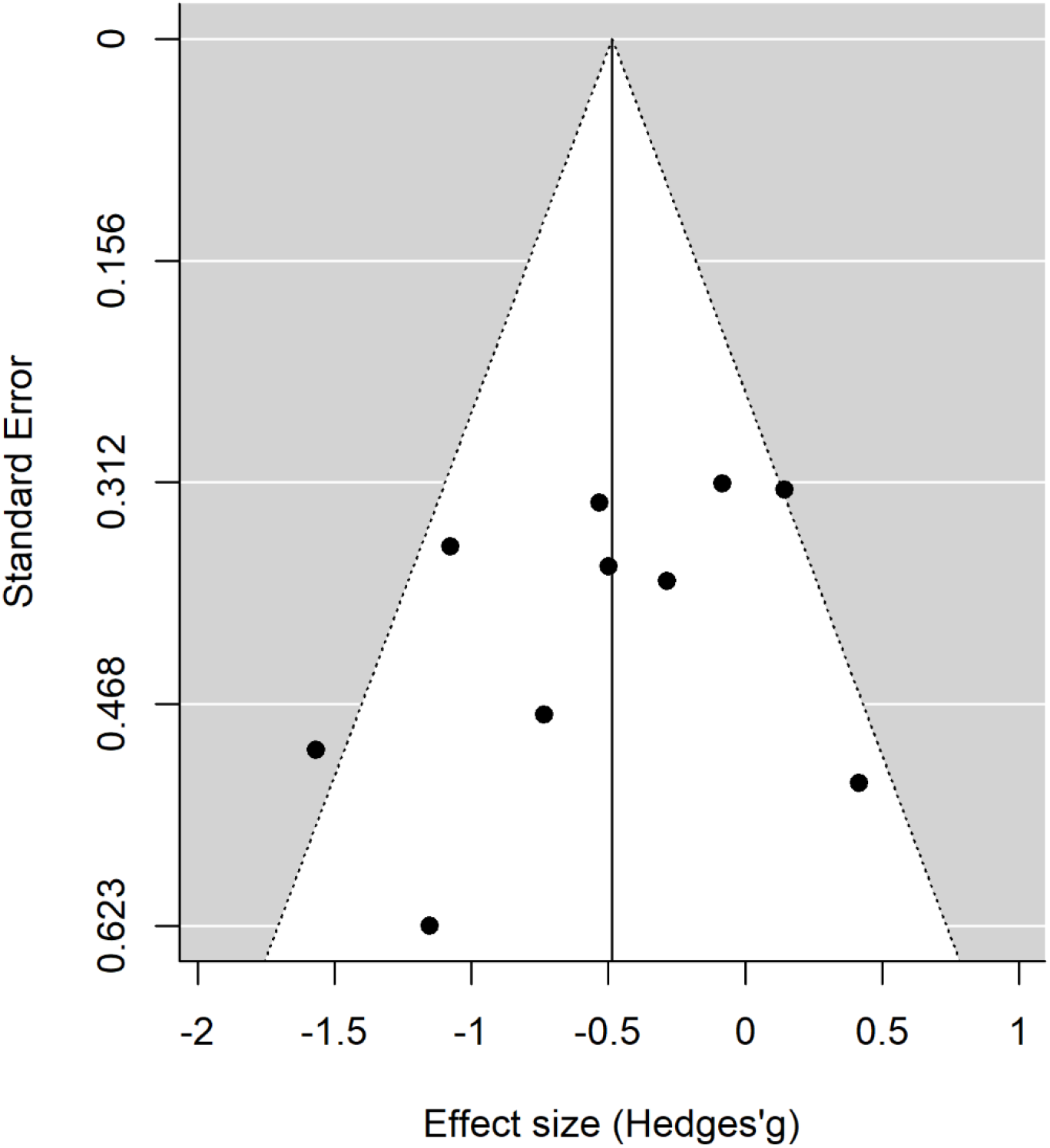
Publication bias assessed by the funnel plot.

#### General anxiety indexes

Since nine of the eleven included studies reported, alongside with the specific anxiety measures, the pre-post real and sham general BAI and HAM-A scores, we run a separate meta-analysis on these scales. As for the specific anxiety symptoms, the random effect model showed a significant medium to the large effect of NIBS protocols in scores reduction as compared to sham treatments (overall SMD of -.81; 95% CI = [-1.45, -.18], p=.012; see Tab 8 for the complete results and Fig. 5 for the forest plot). As for specific symptoms, I^2^ and Q-statistic suggest a high heterogeneity across studies, and Baujat plot suggests study 3 ^90^ as the main source of variance (see Fig. 6). The influence test indeed highlighted this study as an outlier (see Table 8). Thus, the random effect model was re-run, excluding this study from the pool. The global effect of NIBS over general anxiety scores reduction remained significant (overall SMD= -.57; 95% CI = [-1.06, -.07], p=.024; see Table 8 for the complete results). No other study resulted in a significant outlier. We thus proceeded with the moderation analysis with the original set of 9 studies. The inclusion of moderators in the model was not significant (ps> .192, see Table 9). Funnel plot asymmetry (Fig. 7) tested non-significant for both the Egger’s regression test (z = −0.31, p= 0.76) and the Rank correlation analysis (Kendall’s tau = −0.17; p= 0.61).

**Figure 5.**
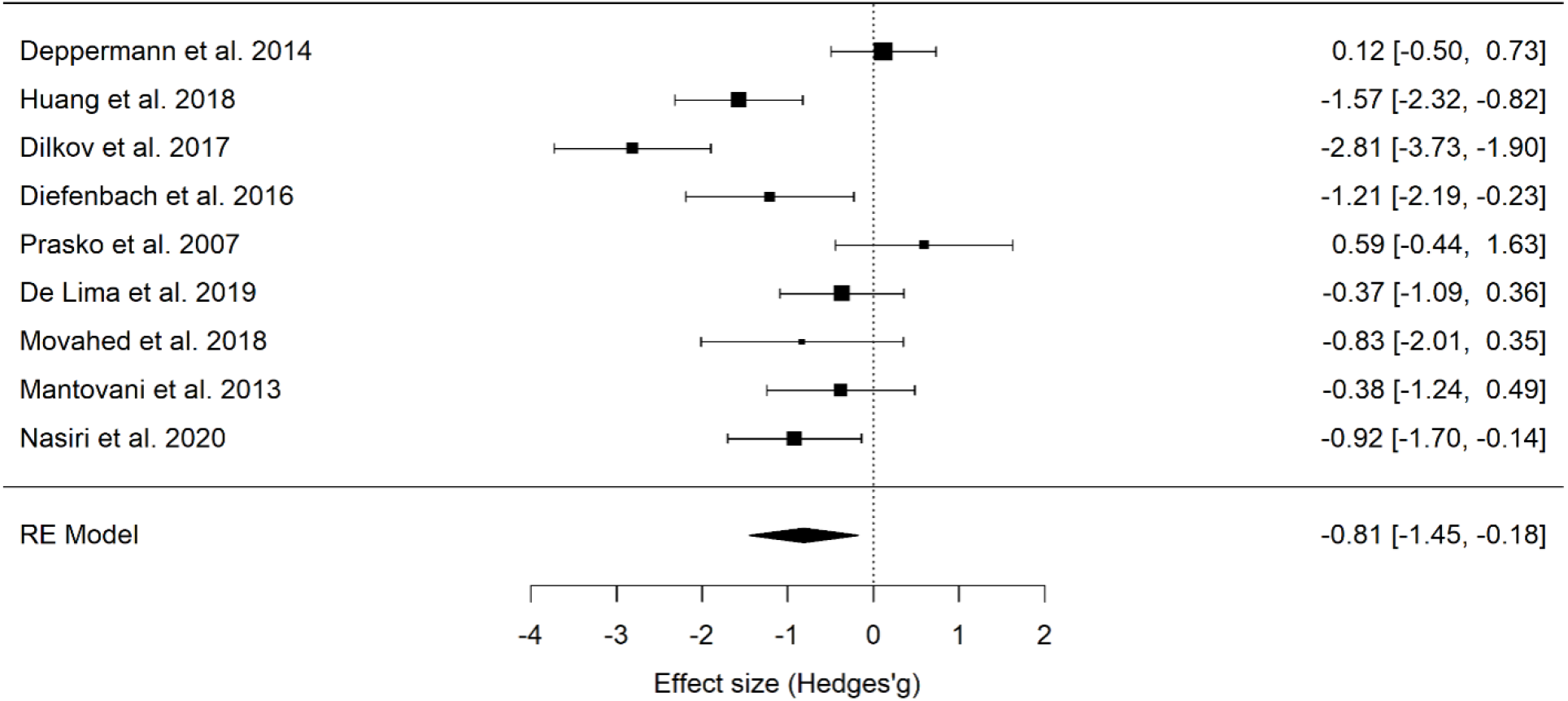
Forest plot of the effect size of NIBS on continuous general anxiety questionnaire scores.

**Figure 6.**
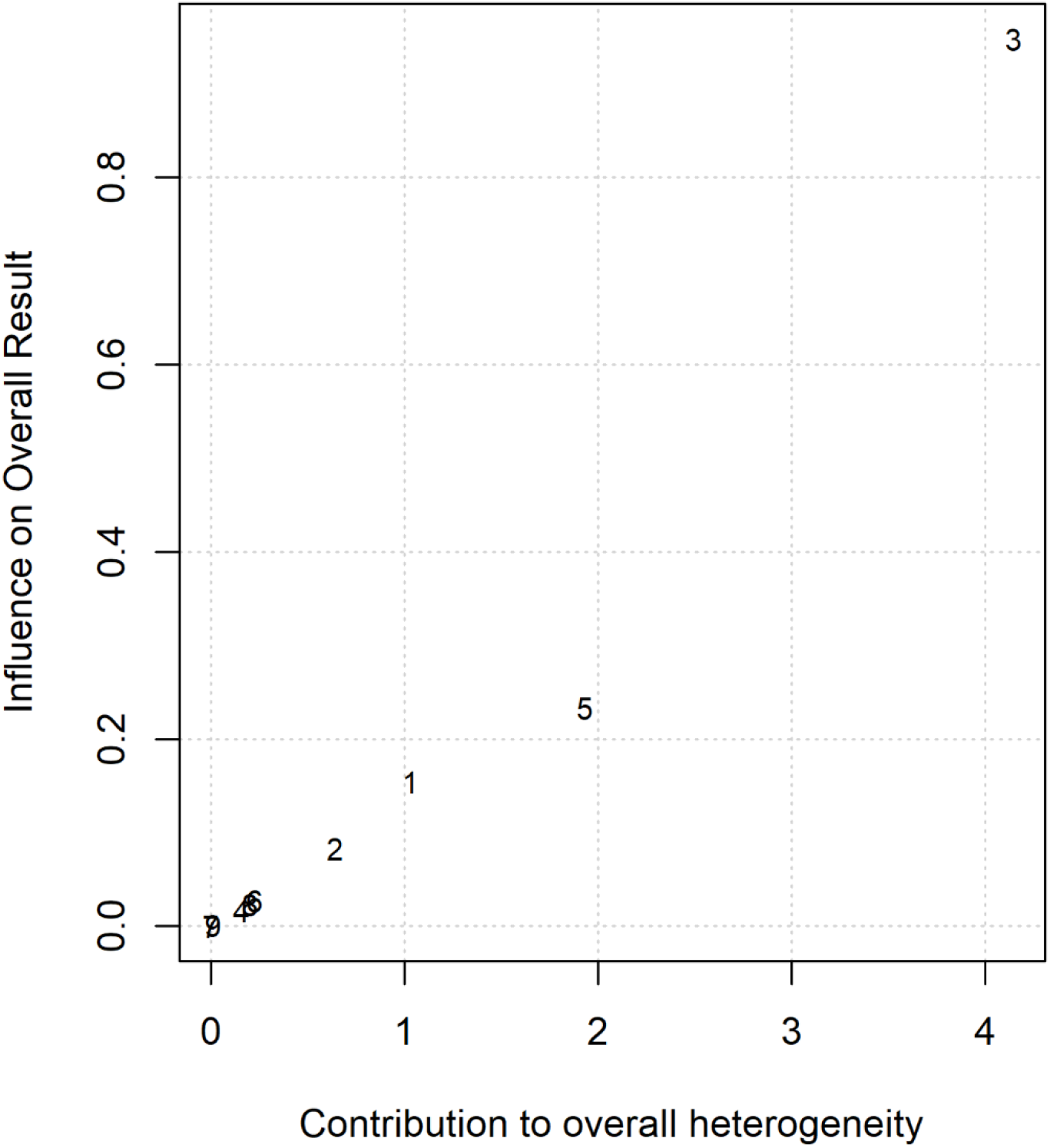
Baujat plot of studies distribution in terms of heterogeneity. At a visual inspection, study 3 (Dilkov et al., 2017) seems to contribute to the statistical heterogeneity in the analyzed studies.

**Figure 7.**
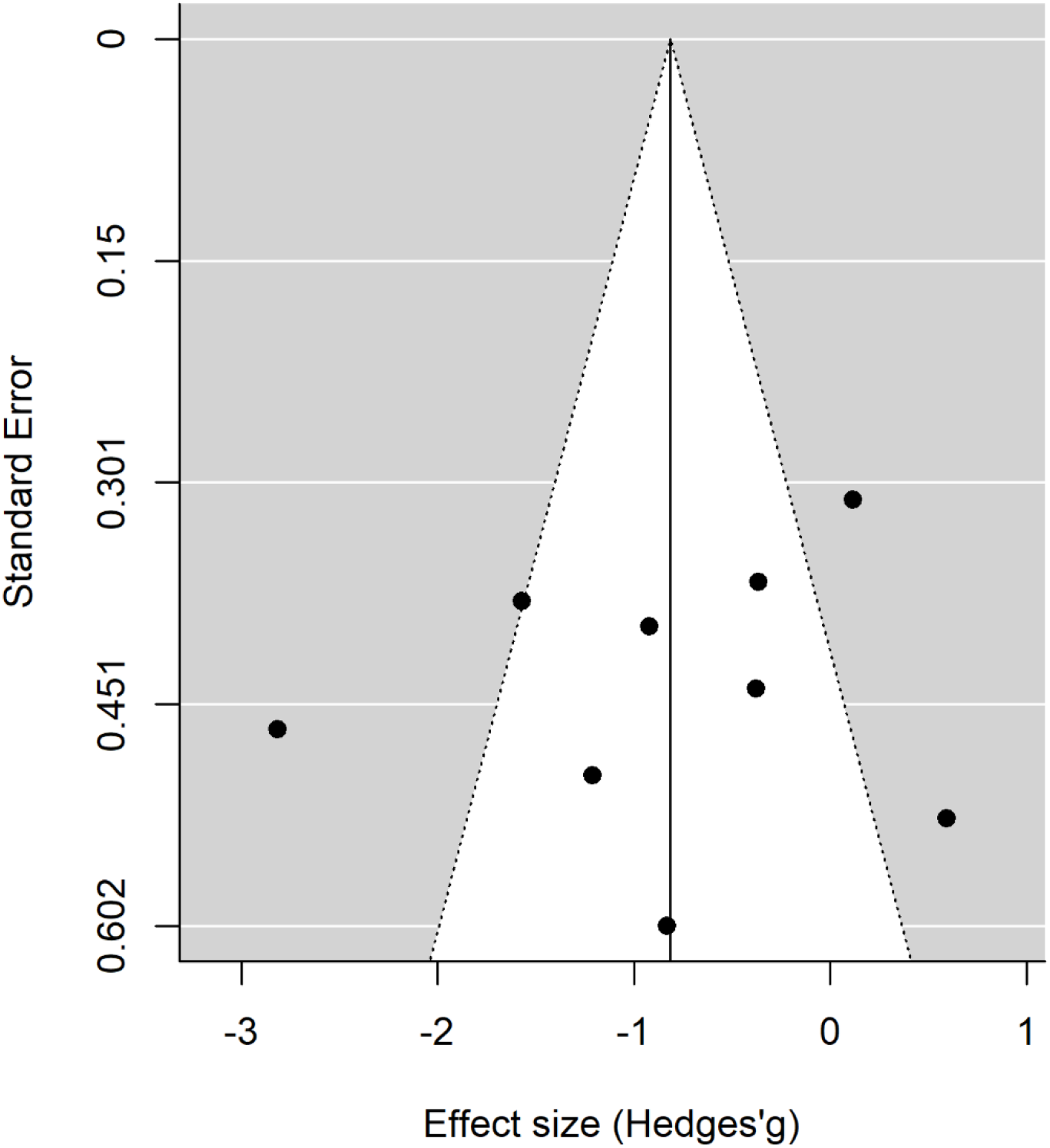
Publication bias assessed by the funnel plot concerning continuous general anxiety questionnaire scores.

#### Depression scales

Seven of the final pool studies reported depression scale scores (see Table 1) both before and after the intervention. The random effect model reported a significant global effect of NIBS in reducing the scores at the depression inventories compared to sham interventions (overall SMD= −.98; 95% CI = [-1.62, −.35], p= .002; see Tab 8 for the complete results and Fig. 8 for the forest plot). I^2^and Q-statistic suggest a high heterogeneity across studies. The Baujat plot suggests study 3 ^90^ as the main variance source. The influence test identified this study as an outlier (see Table 8). Even excluding this study from the meta-analysis, the model highlighted a significant effect of NIBS on depression scores reduction (overall SMD= −.64; 95% CI= [-0.98, −0.31], p<.001; see Tab 8 for the complete results). No further study resulted in an outlier from the influence analysis. We thus proceeded with the moderation analysis with the original set of 7 studies. Analysis of moderators indicated a trend toward significance for the effect of the number of stimulation sessions on depression symptoms reduction (QM _(1)_ = 3.1, p= 0.076), with a higher reduction when the number of sessions increased. No effect was found for the presence of comorbidity in the depression scores after treatment (p=.157). Funnel plot asymmetry (Fig. 9) tested non-significant for both the Egger’s regression test (z = −0.8, p= 0.43) and the Rank correlation analysis (Kendall’s tau = −0.07; p= 1).

**Figure 8.**
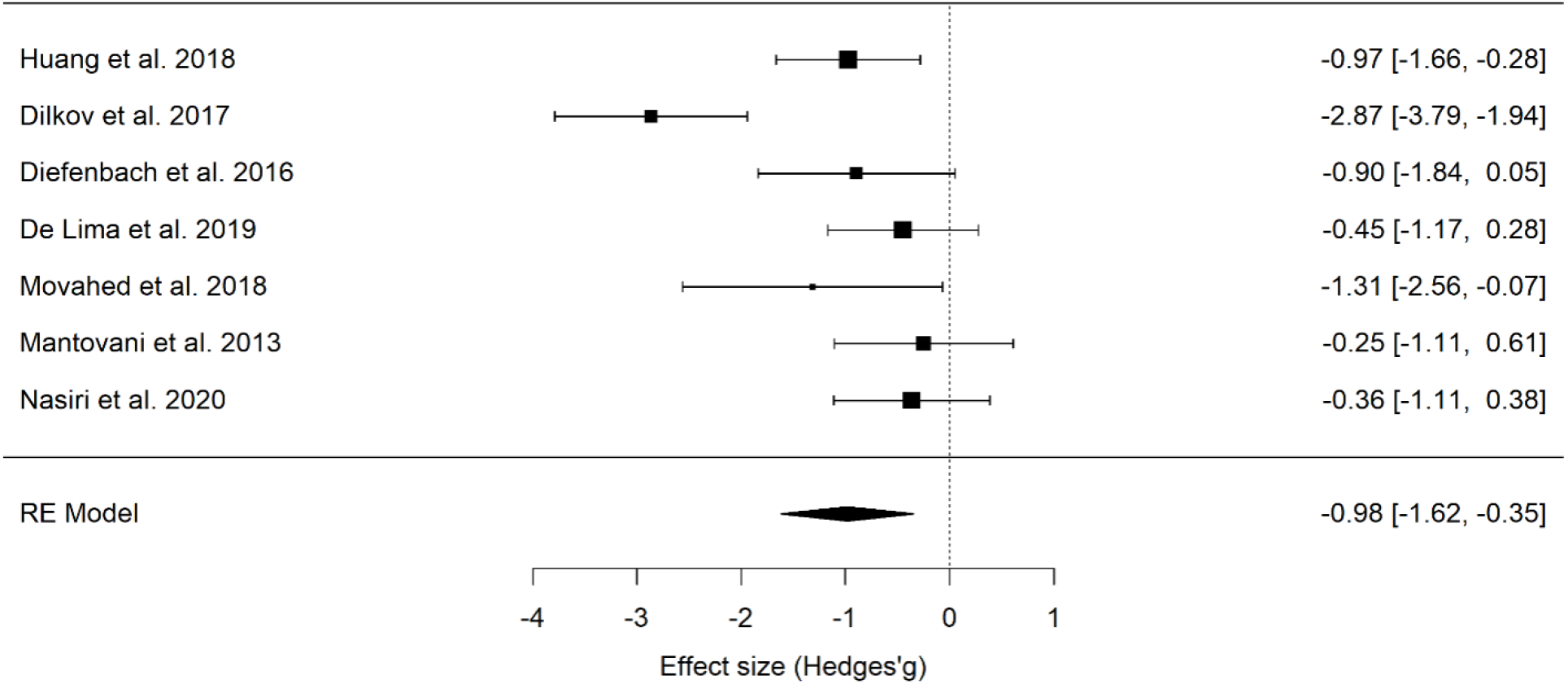
Forest plot of the effect size of NIBS on continuous depression questionnaire scores.

**Figure 9.**
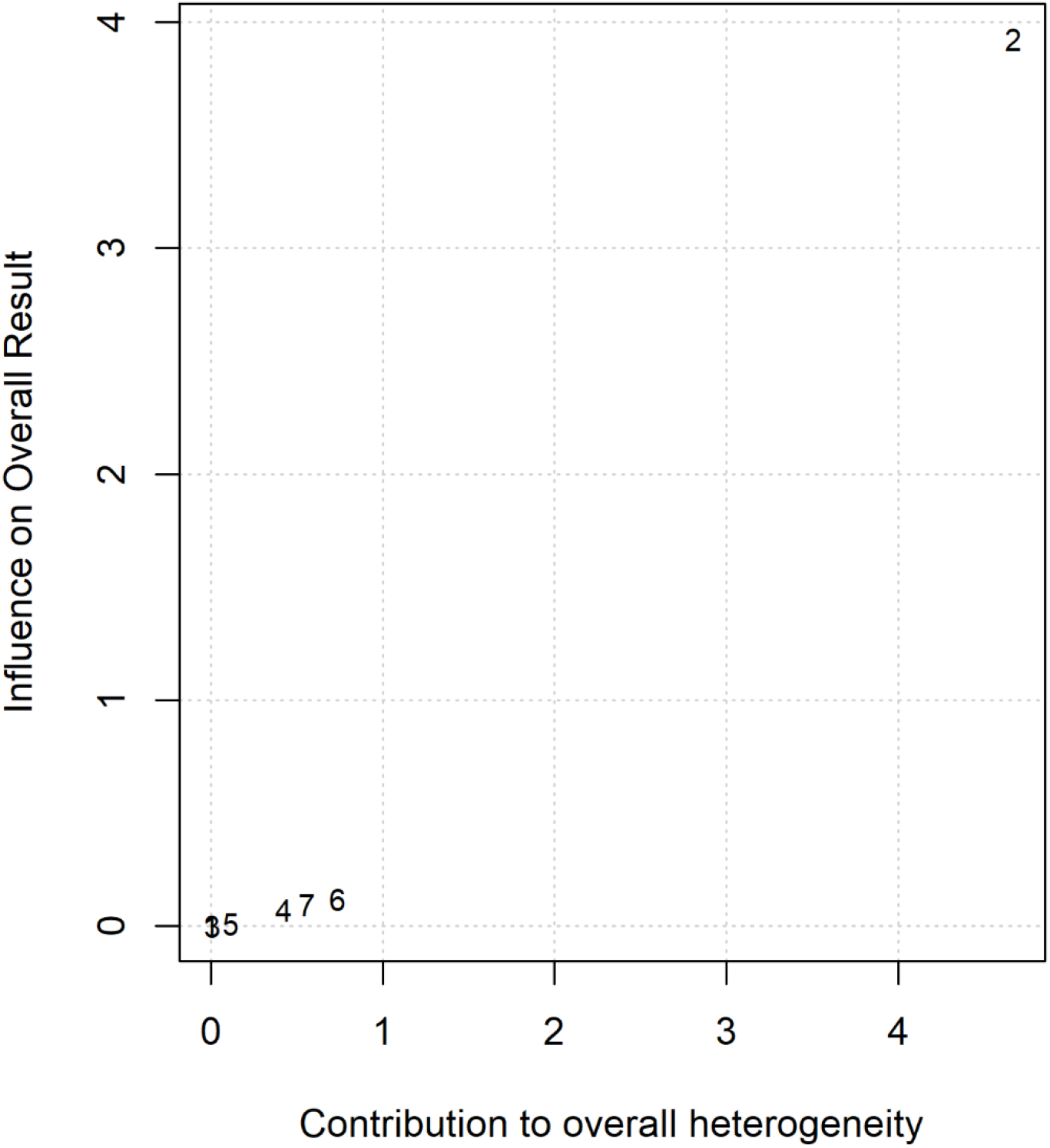
Baujat plot of studies distribution in terms of heterogeneity. At a visual inspection, study 2 (Dilkov et al., 2017) seems to contribute to the statistical heterogeneity in the analyzed studies.

**Figure 10.**
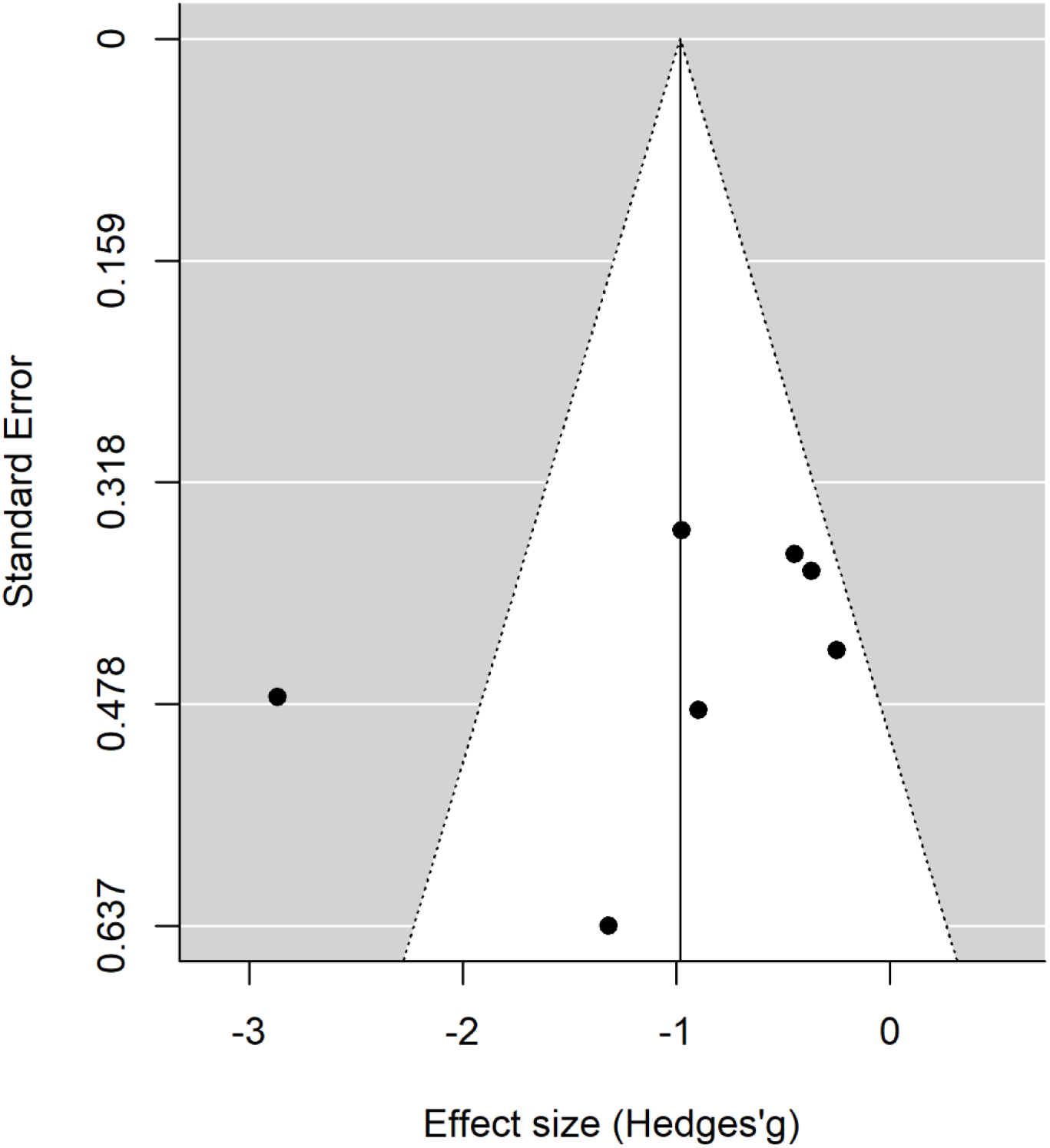
Publication bias assessed by the funnel plot on continuous depression questionnaire scores.

## Discussion

Over the last decades, the debate concerning the high rate of psychiatric patients not responding to the conventional treatment and the low compliance to the pharmacological intervention due to the significant side effects brought to the increasing demand for novel and complementary approaches, among which the application of NIBS.

Despite the effectiveness of rTMS in depression is well-recognized, its clinical use worldwide accepted ^39^, and recent tDCS panel experts’ guidelines points the same direction ^65^, to date no evidence for efficacy have been suggested yet for anxiety disorders ^64–66, 139^, due to the low number of studies specifically investigating this topic.

Therefore, in the present work, we systematically reviewed and quantitatively analyzed the effectiveness of NIBS in ameliorating the clinical symptoms of anxiety disorders. Peer-review, English-written original studies were included in the present work. Given the importance of comparing real stimulation with a placebo or control treatment, we only included studies that randomized participants in these two categories, excluding studies reporting only real stimulation conditions.

Overall, eleven articles were included following the highlighted criteria. Studies differed for the specific disease in the anxiety spectrum: ten out of eleven studies (all but ^90^), indeed, reported questionnaires investigating specific anxiety disease symptoms (e.g., the PDSS in panic disorder). Additionally, nine out of eleven studies (all but ^91, 124^) included a general anxiety measure, namely the HAM-A or the BAI. Therefore, we decided to run two separated meta-analyses concerning anxiety symptoms. The first included for each paper a specific-disorder questionnaire, which should be the specific target of each treatment approach. The second one included a general anxiety questionnaire, namely the HAM-A or the BAI; in this case, the clinician-administered HAM-A was preferred when available. Interestingly, seven papers ^91, 118, 123, 124^ also included scores at a depression scale (HAM-D or BDI), which was the focus of our third meta-analysis. Indeed, it is well-known that anxiety and depression disorders are often in comorbidity and share some commonalities in the neural substrate involved. Therefore, we wanted to investigate whether NIBS could be useful in reducing depression symptoms.

Our findings highlighted a significant medium effect of the real stimulation in decreasing anxiety scores compared to the control condition, thus suggesting that NIBS can be useful in reducing anxious symptoms in patients. This effect was significant for both the disorder specific and general anxiety measures, which is in line with the high correlation found between the two measures of anxiety (0.6) and might be due to changes in symptoms that are shared by the different anxiety disorders. Crucially, the effect was not likely influenced by publication or reporting bias. In line with previous systematic reviews ^34^ and meta-analyses ^71, 72^, we also acknowledge the limitation of the present results, which are based on a restricted sample of studies, but a relatively big pool of patients (318 in total).

Concerning the moderation analysis, we included only representative moderators in our analyses, due the small number of studies included. For example, only two studies did not target the DLPFC, preferring the right parietal region PPC ^121^ and the vmPFC ^124^. Therefore, the moderation analysis concerning the target region was run only comparing left vs. right DLPFC stimulation. The analysis of moderators did not highlight significant predictors, possibly due to the limited number of available studies. Only the number of stimulation sessions revealed a trend to significance, with depressive symptoms decreasing for studies including more sessions, in line with a recent meta-analysis ^140^. The influence of NIBS sessions number in modulating depressive symptoms is debated but still controversial. Some studies and meta-analyses reported a non-significant effect of dosage on symptoms modulation ^141, 142^, while others suggest that at least 20-30 sessions (or more) are required for optimal effects ^143, 144^.

Concerning the statistical variability, Q-statistic and I^2^ suggest a high heterogeneity across studies, probably due to the methodological differences across the selected works. Indeed, protocols varied considering participants’ diagnosis and treatment, the inclusion of associated therapies, as well as protocols’ specific parameters, the target brain regions, and intervention duration. Specifically, participants’ diagnosis included generalized anxiety disorder - also combined with insomnia or major depression -, panic disorder with or without agoraphobia – sometimes in comorbidity with major depression -, specific spider phobia. Heterogeneity also characterized the possibility of participants to combine a medication or psychological treatment. Indeed, in three studies ^91, 117, 120^ participants were not allowed to follow a drug therapy, while in others they could continue their treatment or follow a new one ^118^. In the latter situation, a time interval was established before to start the NIBS treatment, which varied across studies but was three weeks at minimum. As for the psychological treatment, only four protocols ^91, 118, 120, 124^ included a psychological intervention. Crucially, psychological and stimulation interventions were not always combined in the same sessions or, in other words, they were not sequentially or simultaneously time locked ^40^. Indeed, in Deppermann et al. ^118^ participants took part in three group sessions of panic disorder psychoeducation, separately from NIBS intervention. In Nasiri et al. ^120^ tDCS was applied in the last two weeks of an emotional disorder psychological treatment (UP), but authors did not specify whether tDCS was applied before, during or after the treatment. Only Notzon and colleagues ^91^ and Herrmann et al. ^124^, provided a combined approach to the specific phobia, delivering a single session of iTBS before the virtual reality exposure. While Notzon and colleagues did not report changes due to the single session intervention, Herrmann and collaborators two-session treatment reported a reduction in anxiety symptoms scores. As previously highlighted by other researchers (see ^40^ for a recent review), the lack of combination between behavioral or cognitive interventions with NIBS is a gap in neuropsychiatric literature research. Indeed, it is well-known that NIBS effects are known to be state-dependent, meaning that the state of the stimulated regions during NIBS administration has a great influence on the effects of stimulation on cortical excitability ^56, 145–147^ and behavior ^148–150^. Moreover, converging evidence suggested that both stimulation and psychotherapy can modulate brain connectivity ^151, 152^, thus suggesting the importance of time-locking brain stimulation and behavioral engagement to investigate the possibility of maximizing their effects. A similar approach has been initially applied with stroke patients in the neuro-rehabilitation field, combining NIBS with motor and speech training (for recent reviews see ^153, 154^). In neuropsychiatric disorders, combined interventions are still in their infancy ^40, 155^, even in treating depression that received stronger attention by researchers ^40, 142^. Concerning anxiety disorders, for example, Heeren and colleagues ^115^ combined the Attentional Bias Modification technique with anodal and sham tDCS to reduce the bias for threat in patients diagnosed with social anxiety. The study had a crossover design, indeed participants performed only two sessions: a sham and a real one, and authors reported a significant reduction in the bias in the real vs. sham stimulation. As for other disorders, Segrave and colleagues ^156^ combined tDCS with a simultaneous cognitive treatment (cognitive control therapy) in patients with major depression disorders in a five consecutive daily session treatment. Real and sham tDCS equally improved depression symptoms after the fifth session of the protocol; however, effects were maintained at the three-weeks follow up only for the group assigned to the real stimulation. In schizophrenic patients, brain stimulation has been combined with cognitive remediation, trying to improve the cognitive deficits typical of the disease, but produced mixed results ^157^. In conclusion, there is evidence from experimental, behavioral, and clinical research suggesting that the coupling of NIBS with a concomitant treatment might enhance the efficacy of each intervention alone. However, results are scarce and controversial, and the topic needs further investigation to sustain this claim.

As for the included NIBS techniques, most studies included a TMS intervention, being either rTMS ^90, 119, 121–124^ or iTBS ^91, 118^. Only three studies ^89, 117, 120^ were based on a tDCS intervention. This choice is in line with knowledge concerning depression treatment, in which rTMS is considered a useful method to treat drug resistant depression and with the idea that rTMS has stronger spatial resolution as compared to tDCS ^158^. In a combined approach perspective, however, tDCS can be a convenient option, by reducing exogenous “distractions” due to the rTMS-induced noise and muscular contractions. The latter can be annoying or painful, especially when applied to the prefrontal regions, which are the one typically targeted in the revised treatments.

Recently, besides tDCS and rTMS, deep TMS gained ground in treating OCD ^159, 160^ and MDD symptoms (see ^161^ for a meta-analysis), receiving FDA clearance for both treatments. This technique used the same principles of TMS but delivers current through a specially designed H-coil, that can modulate cortical excitability up to 6 cm of depth, therefore reaching not only cerebral cortex activity but also the activity of deeper neural circuits ^162^. To our knowledge, no previous studies investigate deep TMS application to anxiety disorders, and no articles concerning this technique came from our literature search combining “transcranial magnetic stimulation” or “TMS” with the five anxiety categories. However, considering that “deep TMS” was not systematically searched, we combined the keywords “deep TMS” with each of the anxiety disorders in the three previously investigated databases. Pubmed and Scopus research reported no results, while Web of Science search produced three results^4^: a non-original study ^163^, one including only animals ^164^, one involving depressed patients ^165^. The lack of studies combining deep TMS with anxiety disorders reflects the general limited number of studies investigating NIBS and anxiety disorders as compared to other psychiatric conditions, thus highlighting the importance of shedding light in this field.

Concerning the clinical comorbidity of anxiety and depressive disorders, our results highlighted the efficacy of NIBS in reducing depression scores as compared to the control condition, an effect which was not merely trained by studies in which comorbidity was formally diagnosed in the tested sample. This finding is in line with previous studies that investigated rTMS effectiveness in reducing anxiety symptoms during the treatment of depressed patients. In one of the largest studies, Chen and colleagues ^166^ investigated the efficacy of left-DLPFC high-frequency, right-DLPFC low-frequency and sequential bilateral rTMS (i.e., high-frequency left-followed by low-frequency right DLPFC) in a sample of 697 participants. The stimulation protocols showed an overall efficacy of the three protocols in reducing anxiety and depressive symptoms, without indicating a stronger therapeutic effectiveness of one more than other treatment. In another study, Clarke and colleagues ^104^, analyzed data on a sample of 248 patients with treatment resistant depression, of which 172 had one or more comorbid anxiety disorders. rTMS was applied using 1 Hz to the right DLPFC or a sequential bilateral protocol (10 Hz over the left DLPFC and 1 HZ over the right DLPFC). Interestingly, rTMS reduced anxiety levels in patients with and without a formal anxiety diagnosis, as shown by the significant reduction of HAM-A scores in both sub-groups. Similarly, in our sample nine out of eleven interventions targeted the left or right DLPFC, while only two studies ^121, 124^ targeted a different site, namely the right PPC and the vmPFC, respectively. Crucially, when applied over the right hemisphere (DLPFC or PPC) stimulation was inhibitory (with the only exception of ^90^, who applied an excitatory protocol over the rDLPFC), with cathodal tDCS or low frequency (1 Hz) rTMS application, while over the left DLPFC, all studies applied excitatory protocols, as iTBS and anodal tDCS. This choice is in line with previous knowledge concerning neural underpinning of anxiety disorders, which suggests the left DLPFC is typically hypoactive in anxiety disorders, while right DLPFC seems too hyperactive ^32, 33, 167^. The overlap between the targeted regions and inhibition/excitation protocols explains the reported efficacy of NIBS in reducing both anxiety and depression scores compared to the control conditions. Indeed, despite international guidelines and FDA approval recommend the application of excitatory (high frequency rTMS / deep TMS or anodal tDCS) over the left DLPFC, it is known that NIBS can influence brain excitability also through interhemispheric projections. According to this idea, a change in excitability in one hemisphere, also induced by an exogenous stimulation as NIBS delivery, might induce indirect changes in the other hemisphere excitability and eventually in the behavioral outcome. Such effect has been previously reported for cognitive and motor tasks involving prefrontal and frontal regions ^168–170^ and neurorehabilitation, especially involving post-stroke patients ^171, 172^.

This result is exciting and paves the way to a future avenue to specifically investigate the phenomenological overlapping of depression and anxiety disorders. Indeed, not only the stimulation of a similar brain network modulates both anxiety and depression symptoms, but also some antidepressant drugs – primarily serotonin/adrenaline reuptake inhibitors show an effectiveness in treating both disorders, suggesting a similarity also concerning the neurochemical basis of the two syndromes. A recent study by Maggioni and colleagues ^173^ specifically investigated neural commonalities and differences between anxiety and depression by using structural magnetic resonance imaging. Albeit preliminary, authors’ findings suggested that the clinical similarities between major depression and anxiety might rely on common prefrontal alterations, involving left orbitofrontal thinning, while frontotemporal abnormalities are traceable in major depression disorder and parietal are specific to panic and social anxiety disorders.

It is interesting to highlight that prefrontal regions are generally linked to emotional processing and regulation ^30, 174–176^, which are known to be at the basis of both anxiety and depression development and maintenance.

For instance, previous studies with healthy participants suggested that left DLPFC stimulation has positive effects in modulating several cognitive, emotional, and neural processes which are relevant to anxiety ^28, 115, 177, 178^, see ^179^ for a systematic revision of tDCS effects in anxiety disorders or anxious behaviors in healthy participants

A last comment should be done on the outcome measures. In the included papers, ten out of eleven studies used as outcome measure scores at validated explicit questionnaires investigating physical and psychological anxiety and depressive symptoms. Only one study ^91^, investigated psychophysiological measures in addition to questionnaires, namely skin conductance level (SCL) and heart rate variability (HRV). Authors did not report differences in SCL but found a modulation in HRV in real vs. sham iTBS, which was independent from participants’ sample (patients vs. healthy individuals). To date, it is not usual to measure implicit psychophysiological measures as indicators of treatment effectiveness when applying NIBS ^34^. However, they might be an index not only to assess treatment improvements, but also to “dose” the intervention in a flexible way and use these measures as a predictor of treatment outcome. For example, in a previous study with Veterans affected by PTSD, baseline startle response to virtual reality combat-related scenes was predictive of the clinical outcome, with higher startle responses predicting greater changes in symptoms severity at the end of the 6 weeks treatment ^180^.

To conclude, research investigating the relationship between NIBS and anxiety disorders is still in an embryonal state. Overall, only few studies targeting patients with anxiety disorders are available: many authors, indeed, focused on healthy participants with a high trait of anxiety. Concerning the studies including a clinical sample, only few protocols investigated NIBS efficacy at a group-level. Moreover, the inclusion of a sham or control condition to be compared to the real stimulation is not the standard in researchers’ procedure, despite the placebo effect of NIBS techniques is well-known, in both participants and experimenters, thus highlighting the importance of applying double blind procedures ^181, 182^. Future studies should also move in the direction of coupling NIBS with behavioral / cognitive interventions, investigating whether coupled treatment can be more effective than monotherapies.

Although preliminary, our results suggest that NIBS can be effective in decreasing anxiety and depressive symptoms in anxiety disorders, thereby paving the way for treatment protocols including NIBS. However, further research is necessary to optimize the protocols in terms of duration, location, intensity, technique, also related to potential inter-individual differences in response to neuromodulation induced by these NIBS ^54^.

## Data Availability

Data are available upon reasonable request

## Footnotes

1 Post-traumatic stress disorder and obsessive-compulsive disorders no longer fall in this grouping, and therefore will be not considered in the present meta-analysis.

2 Only one study (Huang et al., 2018) targeted a region different from the left and right DLPFC, therefore we did not include it when analyzing the moderation effect of the target region.

3 The UP is a transdiagnostic protocol treatment of emotional disorders, which aims at targeting common features of anxiety and mood disorders, using a single psychological treatment.

4 The three results came from the combination of “deep TMS” AND “generalized anxiety disorder” ^163^, “deep TMS” AND “specific phobia” ^164^, “deep TMS” AND “social anxiety disorder” ^165^.

